# Overexpression of Cancer- and Neurotransmitter-Associated Genes in the Nucleus Accumbens of Smokers

**DOI:** 10.1101/2022.09.30.22280587

**Authors:** Richard Stein, Firoza Mamdani, Blynn Bunney, Preston Cartagena, Marquis P. Vawter, Alan F. Schatzberg, Jack Barchas, Francis S. Lee, Richard Myers, Stanley J. Watson, Huda Akil, William E. Bunney, Adolfo Sequeira

## Abstract

The effects of smoking in the human brain were explored at the molecular level in the *nucleus accumbens*(NAcc), a key brain region involved in tobacco addiction. Gene expression data from post-mortem NAcc were analyzed according to smoking habits: Never smokers, Former smokers and Current smokers at the time of death. The effect of smoking was determined using an ANCOVA model, controlling for potential confounders (psychiatric diagnosis, gender, age, post-mortem interval, and brain pH) followed by pair-wise post-hoc comparisons. Q-values (false discovery rate adjusted p-values) < 0.05 were used in combination with a fold change of > ±1.3 to identify the most relevant genes. The greatest number of differentially expressed genes (DEGs) were found in subjects with a recent history of smoking (Current smokers) compared to either Former or Never smokers. Only two genes were differentially expressed between Former and Never smokers, suggesting that the effects of smoking on gene expression in the brain may be transient. Ingenuity Pathway Analysis (IPA) of DEGs identified a significant over-representation of neurotransmitter system genes (glutamate, GABA) in Current smokers. IPA also revealed many genes associated with cancer in Current smokers compared to Former and Never smokers despite no known cancer in any subjects. Genes associated with neoplasms, glioblastoma, gliomas and tumor regulations are among the top 10 transcripts.Our findings show that active smokers have a significant increase in cancer-related genes and alterations in glutamate and GABA neurotransmitter systems in the NAcc. To our knowledge this is the first study to identify cancer-related genes in the NAcc in Current smokers who have no evidence of cancer.

## Introduction

Tobacco use disorder affects more than a billion people worldwide (Leone and Evers-Casey 2022) and is considered one of the most preventable public health risks responsible for an estimated 7 million deaths per year (World Health Organization 2021). In the United States, an estimated 30.8 million individuals are smokers, costing an average of $225 billion for treatment of smoking-related diseases (CDC 2022). Importantly, life expectancy of smokers is decreased by 10 years and contributes to an estimated 28% of all cancer deaths in the United States in adults 35 years and older (CDC 2020). Cigarette smoke contains over 7000 chemical carcinogens (Jamal et al. 2021). Despite multiple attempts to stop smoking, only a small percentage succeed(NIDA 2020). This is particularly problematic in psychiatric patients where the incidence of smoking is substantially higher than in the general population (Fornaro et al. 2022). In 2020, the incidence of smoking was 12.5% in adults over the age of 18 years (CDC 2022a)] in contrast to 33.4% in major depressive disorder (MDD), 46.3% in bipolar disorder (BPD) and 65% in schizophrenia (Fornaro et al. 2022). In this article we aim to investigate the molecular effects of cigarette smoking status in the *Nucleus Accumbens* (Nacc), a central area of the brain involved in addiction. A greater understanding of the molecular basis for addiction can provide important insights to developing improved methods to treat and potentially prevent tobacco addiction.

Nicotine is considered the key ingredient in cigarette smoke associated with reward and addiction. In brain, nicotine binds to the nicotinic acetylcholine receptors (nAChRs), and it is interactions with subtypes of the nAChRs that determine its effects on neurotransmitters including glutamate, dopamine, and GABA which influence a variety of downstream affects that initiate and sustain nicotine addiction(Fowler, Turner, and Imad Damaj 2020). In humans, nicotine induces a dose-dependent increase in neuronal activity in the NAcc, amygdala, cingulate and frontal lobe consistent with its behavioral reinforcing effects (Stein et al. 1998). These structures form an integral part of the mesolimbic reward system with dopamine as its primary neurotransmitter. The mesolimbic circuitry originates in the ventral tegmental area (VTA) and projects to the NAcc, prefrontal cortex, and amygdala (Li et al. 2014).In addition to dopaminergic input from the VTA, the NAccalso receives glutamatergic projections from the prefrontal cortex, hippocampus, amygdala and thalamic nuclei(Pistillo et al. 2015). Together with the VTA, the NAcc plays a central role in addiction and reward. Excitatory inputs from N-methyl-D-aspartate (NMDA) receptors in the VTA increase dopamine levels in the NAcc via activation of presynaptic nAChRs on glutamatergic terminals. Nicotine-induced dopamine levels are also modulated by inhibitory GABA-ergic input including GABA-ergic inhibitory afferents to VTA dopaminergic receptors, inhibitory GABA-ergic interneurons within the VTA and inhibitory medium spiny neurons in the NAcc (Kim et al. 2019, Watkins, Koob, and Markou 2000, D’Souza and Markou 2013, Wills et al. 2022, Pistillo et al. 2015).

The effect of nicotine on gene expression in the NAcc is relatively understudied compared to that of other regions in the mesolimbic circuitry. Animal data provide evidence that the NAcc contains ‘nicotine sensitive’ genes that have the capacity for up-or downregulation following acute or chronic exposure (Kane et al. 2004). However, very little is understood about long-term gene expression effects of nicotine in the NAcc in the human brain. Postmortem investigations have been conducted across brain regions, particularly in areas implicated in the addiction neurocircuitry (e.g., cortical, striatal, and limbic regions) (Zillich et al. 2022, Webb et al. 2015, Markunas et al. 2021, Sey et al. 2022, Quach et al. 2020, Barrie et al. 2017). However, relatively few studies used RNA-seq to analyze the NAcc, a key area associated with addiction and a target of deep brain stimulation (DBS) for treatment-resistant substance abuse including tobacco (Navarro et al. 2022, Chang et al. 2022). Flatscher-Bader et al.(Flatscher-Bader et al. 2010), performed microarray analyses in the human NAcc and VTA in subjects with comorbidalcohol and tobacco use,non-smoking alcoholics, and smoking non-alcoholics. They identified ‘smoking sensitive’ geneswhich distinguished smokers from non-smokers. Mexal and colleagues performed targeted (qPCR) gene expression (Mexal et al. 2010) and microarray (Mexal et al. 2005) analyses of the effect of smoking in the human hippocampus of controls and patients with schizophrenia.The first genome-wide DNA methylation and transcriptomic study to identify changes associated with smokingin human postmortem NAcc comparing current smokers to non-smokers was undertaken by Markunas and colleagues(Markunas et al. 2021).They identified seven regions of differential methylation associated with smoking;three regions were in the proximity of genes previously identified in blood-based studies, while the remaining four regions are novel findings in the NAcc (ABLIM3, APCDD1L, MTMR6, CTCF)(Markunas et al. 2021).

In this study, we used a transcriptome-wide microarray dataset to investigate the effect of smoking status (Current, Former or Never smokers) on the NAcc of individuals with mood disorders and in psychiatrically healthy controls.Findings show that active smokers have significantly increased alterations in genes related to glutamate and GABA receptor and in cancer-associated genes in the NAcc.

## Materials and Methods

Postmortem NAccsamples (N=51) were obtained from the University of California, Irvine (UCI) Pritzker Brain Bank (UCIPBB) through a uniform process approved by the Institutional Review Board. All subjects went through an extensive review of multiple sources of information including the medical examiner’s conclusions, coroner’s investigation, medical and psychiatric records, and interviews of the decedents’ next-of-kin in order to rule out any neurological disorder and cancer. Additionally, a neuropathological examination of each brain was conducted to exclude cases with cerebrovascular disease (infarcts or hemorrhages), subdural hematoma, brain cancer, or other significant pathological features.

Our psychological autopsy protocol is based largely on procedures validated by Kelly and Mann (Kelly and Mann 1996). Details of death are evaluated in order to assign an Agonal Factor Score to each subject in accordance with previous established guidelines regarding agonal factors (Vawter et al. 2006, Tomita et al. 2004, Preece and Cairns 2003, Li et al. 2004, Hynd et al. 2003, Harrison et al. 1995, Barton et al. 1993). Brains were initially screened for agonal factors at the time of consent to exclude brains from subjects with *ante-mortem* conditions potentially affecting nucleic acids and proteins. The human brain dissection and freezing protocol is described in detail elsewhere (Tomita et al. 2004, Bunney et al. 2003). Freezing is done in an isopentane bath cooled at -40°C and brains are stored in -80°C freezers until dissected and processed. The brain regions were dissected on dry ice using visible landmarks near the regions of interest. The studies make use of de-identified and coded autopsy tissue from the University of California, Irvine Pritzker Brain Bank (UCIPBB) that has a proven track record of providing well-characterized, high-quality material. Brains for this project were carefully selected using criteria that minimize factors affecting tissue integrity such as brain pH, agonal factors and score, agonal duration, post-mortem interval (PMI) and RNA integrity number (RIN) (Vawter et al. 2006, Tomita et al. 2004, Preece and Cairns 2003, Li et al. 2004, Hynd et al. 2003, Harrison et al. 1995, Barton et al. 1993).

Statistical analysis was performed using Partek (St. Louis, MO). Cel files from Affymetrix HG-U133 Plus2.0 chips were processed using GCRMA algorithm, with GCRMA background correction, quantile normalization on Log2 median polish probe set summarization (Irizarry, Wu, and Jaffee 2006, Wu and Irizarry 2004, Irizarry et al. 2003, Bolstad et al. 2003). Outlier detection, identification of problematic arrays, was done using a combination of principal component analysis (PCA), and RNAdeg Slopes. ANCOVA identifiedDEGs between the groups (Current, Former and Never) controlling for diagnosis, age, gender, RIN, and pH. Initial filtering selected the genes that were reliably detected by the microarrays (average log2 data > 5) in at least one of the three groups. After the ANCOVA, post-hoc pair-wise comparisons compared Current to Former, Current to Never and Former to Never smokers. False discovery rate (FDR) corrected for multiple comparisons (Benjamini and Hochberg 1995). Genes were considered differentially expressed if corrected q-value were less than 0.05 and the fold change (FC) higher than ±1.3. The q-value is a p-value after controlling for the false discovery rate (FDR), thus the q-value is a corrected p-value representing the proportion of false positives among all positive results. Pathway analysis was performed using Ingenuity (Redwood City, CA) and the differentially expressed Affymetrix probe sets/genes in each comparison.Protein-protein interaction networks of DEGs were investigated using STRING (**Supplementary Figures 1 and 2)**. Data from the Allen Human Brain Atlas was used to independently determine expression levels of nicotinic receptors in human NAcc (**Supplementary Figure 3)**.

## Results

The current study was based on microarray data from postmortem NAcc in individuals with known smoking status at the time of death. While no differences in pH and post-mortem interval (PMI) were found between the groups, Current smokers were slightly younger(average age ± standard deviation = 44.2 years ± 13.5) than Former (p=0.032; 55.5years ± 16.5) and Never smokers (p=0.048; 52.3 years ± 11.8) **(Supplementary Table 1)**.There was a greater proportion of females among theNever smokers (p=0.028). Our sample population included mood disorder patients (BPD, MDD) and non-psychiatric subjects **(Supplementary Table 2**). According to our detailed psychological autopsy including medical records and next-of-kin interviews, none of the subjects had a diagnosable cancer.Examination of the tissue by a pathologist confirmed no evidence of cancer in the brain. Because our primary outcome was the effect of smoking on gene expression in the NAcc, we first performed an ANCOVA with the three smoking groups and the possible confounders of age, gender and psychiatric diagnosisbefore performing pair-wise post-hoc comparisons between Current and Former, Current and Never, Former and Never smokers (q<0.05; FC≥±1.3).Current smokershad by far the largest number of DEGs after FDR correction when compared to Former (279) and to Never smokers (105), while only two genes passed correction when Former and Never smokers where compared (**Supplementary Table 3**).**Figures 1 and 2** show the top over-represented genes.**Supplementary Tables4 and 5** list all the significant DEGs per comparison.

**Figure 1:**
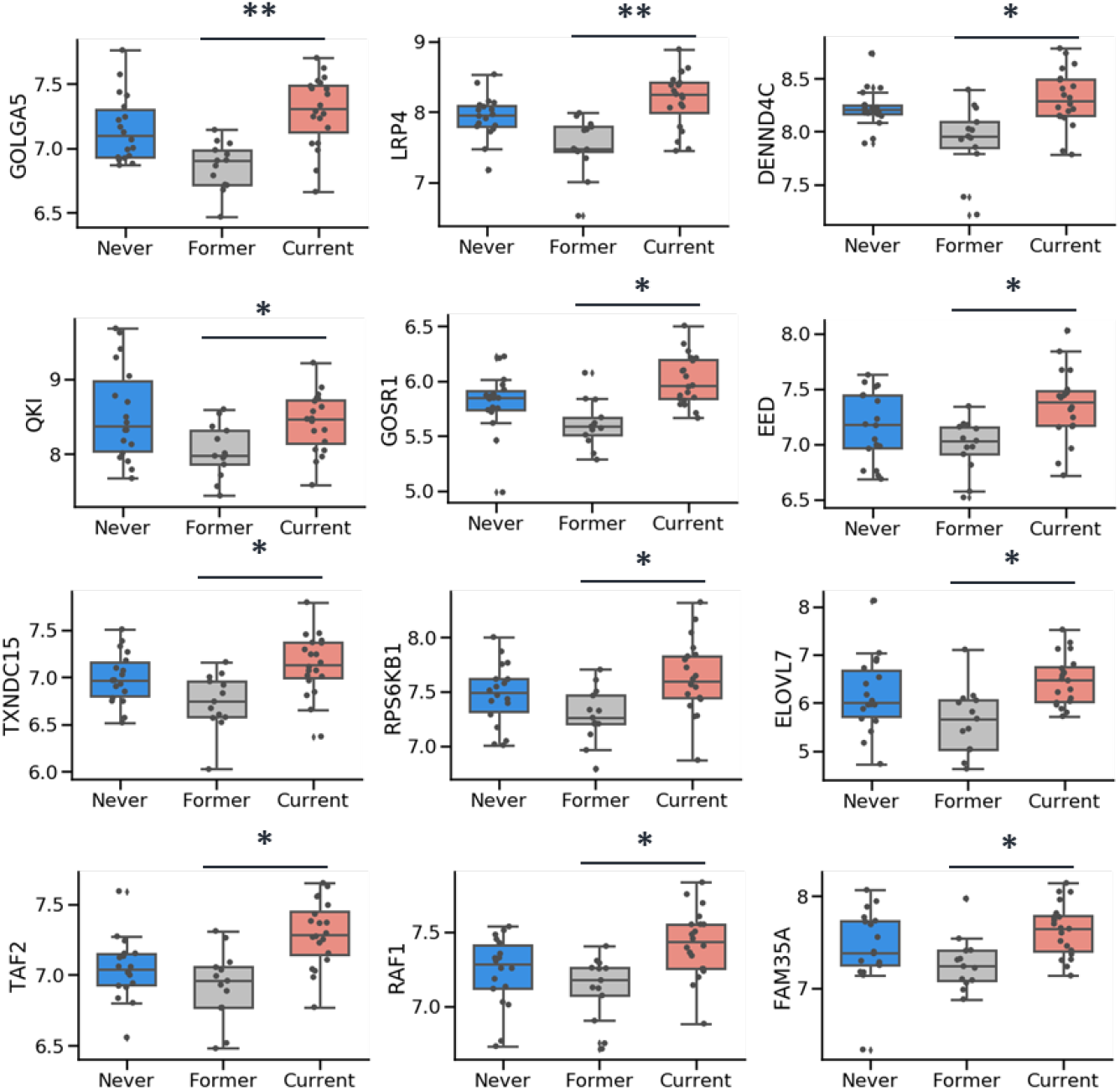
Top differentially expressed genes between Current and Former smokers in the NAcc. After initial ANCOVA was used to identify differentially expressed genes between the groups (Current, Former and Never) with diagnosis, age, gender, RIN, and pH as covariates, post-hoc pair-wise comparisons were conducted comparing Current to Former smokers. Significance required q-value < 0.05 and fold change >±1.3. (*q<0.05, **q<0.01).Significant genes: GOLGA5 (golgin A5), LRP4 (low density lipoprotein receptor-related protein 4), DENND4C (DENN/MADD domain containing 4C), QKI (QKI, KH domain containing, RNA binding), GOSR1 (golgi SNAP receptor complex member 1), EED (embryonic ectoderm development), TXNDC15 (thioredoxin domain containing 15), RPS6KB1 (ribosomal protein S6 kinase, 70kDa, polypeptide 1), ELOVL7 (ELOVL fatty acid elongase 7), TAF2 (TAF2 RNA polymerase II, TATA box binding protein (TBP)-associated factor, 150kDa), RAF1 (v-raf-1 murine leukemia viral oncogene homolog 1), FAM35A (family with sequence similarity 35, member A).

**Figure 2:**
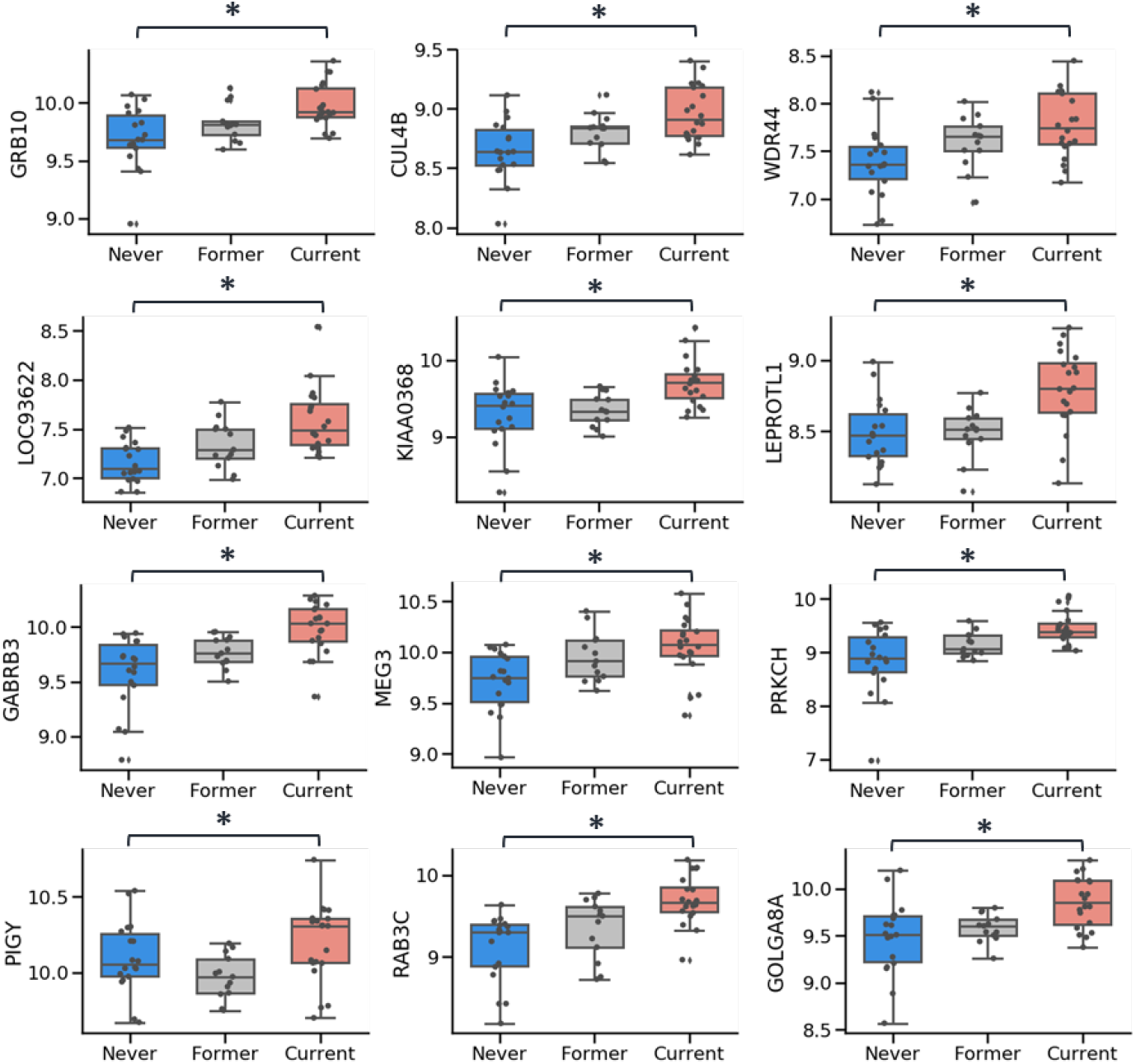
Top differentially expressed genes between Current versus Never smokers in the Nacc. After initial ANCOVA was used to identify differentially expressed genes between the groups (Current, Former and Never) with diagnosis, age, gender, RIN, and pH as covariates, post-hoc pair-wise comparisons were conducted comparing Current to Never smokers. Significance required q-value < 0.05 and fold change >±1.3. (*q<0.05).Significant genes: GRB10 (growth factor receptor-bound protein 10), CUL4B (cullin 4B (Alias KIAA0695)), WDR44 (WD repeat domain 44), LOC93622 (Morf4 family associated protein 1-like 1 pseudogene), KIAA0368 (alias for ECPAS-Ecm29 Proteasome Adaptor And Scaffold), LEPROTL1 (leptin receptor overlapping transcript-like 1), GABRB3 (gamma-aminobutyric acid A receptor, beta 3), MEG3 (maternally expressed 3), PRKCH (protein kinase C, eta), PIGY (phosphatidylinositol glycan anchor biosynthesis, class Y), RAB3C (RAB3C, member RAS oncogene family), GOLGA8A (golgin A8 family, member A).

Current smokers when compared to Former smokers had 265 upregulated genes and 14 downregulated genes out of 279DEGs that survived FDR correction and after controlling for possible confounders (**Figure 1, Supplementary Table 4 and Supplementary Figure 1**).In order to interpret the meaning of these DEGs, we fed the 279 FDR corrected genes into Ingenuity for a core analysis including “canonical pathways” as well as “diseases and disorders” significantly over-represented, as indicated by the p-values and overlap genes in the data and in the pathway for the “Top canonical pathways” and p-value range (as not all genes are known to be involved in any given disease or disorder) and number of molecules or genes for the “Top diseases and disorders.”The top three IPA over-represented canonical pathways**(Table 1)**are related to cancer or are known to be involved in cancer according to the IngenuityKnowledge Base, an extensive repository of biological and chemical “findings” curated from varioussources including databases and the literature: “Regulation of eIF4 and p70S6K Signaling”, “Role of tissue factor in cancer” and “MSP-RON Signaling in Cancer Cells Pathway.” These top pathways represent large mechanistic networks that play a critical role in cancer according to curated data and point to important mechanisms by which smoking can lead to activation of genes known to be involved in cancer despite no evidence of cancer among the subjects. Thus, eIF4 (Eukaryotic Initiation Factor-4) and p70S6K are two protein complexes that play critical roles in translational regulation and in cancer according to IPA. MSP-RON Signaling is a known mechanism used by cancer cells for survival, migration, angiogenesis and chemoresistance. Of the 279 significant genes, 271 are associated with cancer suggesting an overall activation of cancer-related transcripts in Current smokers compared to subjects who stopped smoking (Former Smokers).

**Table 1:** Top pathways, diseases, and disorders over-represented among the differentially expressed genes inCurrent smokers compared to Former smokers.

| Top canonical pathways | p-value | Overlap |
| --- | --- | --- |
| Regulation of eIF4 and p70S6K Signaling | $3.09 \times 10^{-4}$ | 5.0% (9/179) |
| Role of Tissue Factor in Cancer | $4.87 \times 10^{-4}$ | 6.0% (7/116) |
| Adipogenesis pathway | $1.20 \times 10^{-3}$ | 5.2% (7/135) |
| MSP-RON Signaling In Cancer Cells Pathway | $1.36 \times 10^{-3}$ | 5.1% (7/138) |
| Insulin Secretion Signaling Pathway | $1.48 \times 10^{-3}$ | 3.7% (10/268) |
| Top diseases and disorders | p-value range | # Molecules |
| Cancer | $8.63 \times 10^{-3}$ - $1.37 \times 10^{-20}$ | 271 |
| Organismal Injury and Abnormalities | $8.63 \times 10^{-3}$ - $1.37 \times 10^{-20}$ | 272 |
| Gastrointestinal Disease | $8.63 \times 10^{-3}$ - $9.59 \times 10^{-18}$ | 254 |
| Endocrine System Disorders | $7.24 \times 10^{-3}$ - $2.34 \times 10^{-12}$ | 239 |
| Reproductive System Disease | $5.63 \times 10^{-3}$ - $2.36 \times 10^{-10}$ | 205 |

The overlap column shows the number of genes differentially expressed over the total number of genes in the pathway. For diseases and disorders a p-value range of the enrichment is reported from IPA as the exact number of genes involved in any given disease or disorder is not known. eIF4 (Eukaryotic Initiation Factor-4) and p70S6K are two protein complexes that play critical roles in translational regulation and in cancer. MSP-RON Signaling is a known mechanism used by cancer cells for survival, migration, angiogenesis and chemoresistance. Cancer and Organismal Injury and Abnormalities are among the top diseases and disorders dysregulated in Current versus Former smokers, both in fact overlap and point to cancer related genes dysregulation.

Interestingly, two glutamatergic genes were upregulated in Current vs Former smokers, a glutamate transporter (solute carrier family 1 member 1 (SLC1A1)) and the AMPA receptor (glutamate ionotropic receptor AMPA type subunit 2 (GRIA2)). Meanwhile, GABA_B_ receptor 2 (GABBR2) was downregulated in Current versus Former smokers. Current versus Former comparisons revealed several genes involved in metabolic changes related to smoking. The top Current versus Former genes are upregulated with no significant differences observed with Never smokers. Many of these genes are metabolic and involved in hypoxia (HIF3A), monocarboxylate (e.g., lactate/beta-hydroxybutyrate) transport (SLC16A1), mitochondrial glutamate transporter (SLC25A18) and Golgi mediated synaptic vesicle fusion (GOSR1).

When Current smokers were compared to Never smokers,101 genes showed increased expression and 4 showed decreased expression levels out of 105 DEGs that survived FDR correction (**Figure 2, Supplementary Table 5 and Supplementary Figure 2**). Among the top canonical pathways, GABA and dopamine signaling as well as synaptic long-term depression were over-represented **(Table 2)**. As wasthe case of the Current versus Former comparison, nearly all (97/105) of these genes were cancer-related as the top diseases/disorders listed “Cancer” and “Organismal Injury and Abnormalities”.

**Table 2:** Top pathways, diseases, and disorders over-represented among the differentially expressed genes inCurrent smokers compared to Never smokers.

| Top canonical pathways | p-value | Overlap |
| --- | --- | --- |
| Androgen Signaling | $9.33 \times 10^{-5}$ | 3.6% (6/169) |
| GABA Receptor Signaling | $2.62 \times 10^{-4}$ | 3.8% (5/131) |
| Dopamine-DARPP32 Feedback in cAMP signaling | $1.14 \times 10^{-3}$ | 2.8% (5/181) |
| Assembly of RNA Polymerase II Complex | $1.32 \times 10^{-3}$ | 6.0% (3/50) |
| Synaptic Long-Term Depression | $1.55 \times 10^{-3}$ | 2.6% (5/194) |
| Top diseases and disorders | p-value range | # Molecules |
| Cancer | $1.29 \times 10^{-2}$-$7.62 \times 10^{-7}$ | 97 |
| Organismal Injury and Abnormalities | $1.29 \times 10^{-2}$-$7.62 \times 10^{-7}$ | 98 |
| Neurological Disease | $1.29 \times 10^{-2}$-$1.20 \times 10^{-6}$ | 56 |
| Gastrointestinal Disease | $1.29 \times 10^{-2}$-$1.85 \times 10^{-6}$ | 93 |
| Endocrine System Disorders | $7.30 \times 10^{-2}$-$3.14 \times 10^{-6}$ | 83 |

The overlap column shows the number of genes differentially expressed over the total number of genes in the pathway. For diseases and disorders a p-value range of the enrichment is reported from IPA as the exact number of genes involved in any given disease or disorder is not known. Cancer and Organismal Injury and Abnormalities are among the top diseases and disorders dysregulated in Current versus Never smokers, both in fact overlap and point to cancer related genes dysregulation. The androgen signaling pathway is overrepresented in Current vs. Never smoker analysis where males outnumbered females but not in the Current vs. Former analysis where there was no gender difference.

As suggested by the canonical pathway analysis many GABA, glutamateand dopamine related genes were also differentially expressed in Current versus Never smokers**(Figure 3)**. Gamma 2 (gamma-aminobutyric acid type A receptor subunit gamma2 (GABRG2)) and Beta 3 (gamma-aminobutyric acid type A receptor subunit beta3 (GABRB3)) receptors of the GABA_A_ receptor family as well as glutamate decarboxylase 2 (GAD2) involved in GABA production, were upregulated. Two voltage dependent calcium channels, CACNA1E (calcium voltage-gated channel subunit alpha1 E) and CACNA2D3 (calcium voltage-gated channel auxiliary subunit alpha2delta 3), involved in downstream GABA neurotransmission, were similarly upregulated in Current versus Never smokers. Similarly, five genes involved in dopamine neurotransmission were differentially expressed in the NAcc: CACNA1E, CACNA2D3, CSNK1G1, PDIA3, PRKCH.

**Figure 3:**
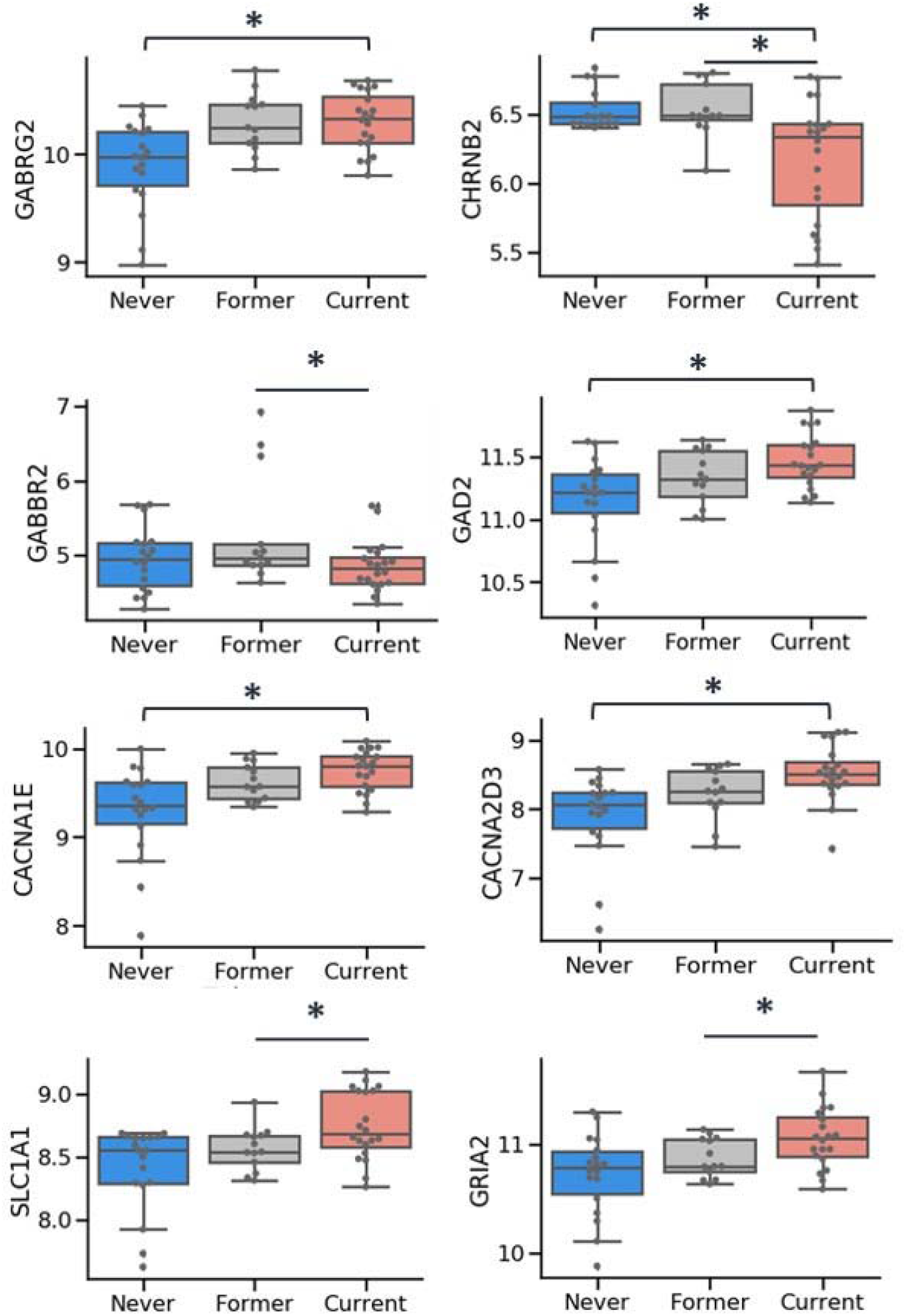
Smoking induced GABA and glutamate gene expression changes in the NAcc. After initial ANCOVA was used to identify differentially expressed genes between the groups (Current, Former and Never) with diagnosis, age, gender, RIN, and pH as covariates, post-hoc pair-wise comparisons were conducted. Significance required q-value < 0.05 and fold change >±1.3. (*q<0.05, **q<0.01).Significant genes: GABRG2, (gamma-aminobutyric acid A receptor, gamma 2), CHRNB2 (Cholinergic Receptor Nicotinic Beta 2 Subunit), GABBR2 (gamma-aminobutyric acid B receptor, 2), GAD2 (Glutamate Decarboxylase 2), CACNA1E (Calcium Voltage-Gated Channel Subunit Alpha1 E), CACNA2D3 (Calcium Voltage-Gated Channel Auxiliary Subunit Alpha2delta 3), SLC1A1 (Solute Carrier Family 1 Member 1), GRIA2 (Glutamate Ionotropic Receptor AMPA Type Subunit 2).

Only 2 genes (AGT and FBXL7) were differentially expressed between Former and Never smokers suggesting that the main driver of gene expression change in the NAcc is the Current smoker status at the time of death. AGT is angiotensinogen (serpin peptidase inhibitor, clade A, member 8), and the associated protein is involved in maintaining blood pressure, body fluid and electrolyte homeostasis. FBXL7 is the F-box and leucine-rich repeat protein 7.

A notable portion of the top DEGs in Current smokers,when compared to Former and to Never smokers, are associated with cancer (**Table 3 and 4** and **Supplemental Tables 2 and 3**) and a good proportion of them have previously been found to be associated with smoking in expression or methylation studies in the diverse tissues obtained from smokers and including buccal swabs, blood and brain tissue.Of the top 10 genes in the Current/Never analysis,five have known associations with cancer: CUL4B (cullin 4B) associated with solid neoplasms(Li and Wang 2017),PRKCH (protein kinase C eta) associated with glioblastoma(Martin and Hussaini 2005), MEG3 (maternally expressed 3) thought to be involved in tumor cell suppression(Al-Rugeebah, Alanazi, and Parine 2019), RABC3 (RAS-associated protein RAB3C) with RAS oncogene familyand gliomas (Raffaniello 2021), and UTP18 (UTP18 Small Subunit Processome Component, over-expressed in cancer(Yang et al. 2015). Lower expression (Current/Never analysis) of GALNTL2/GALNT15 (Polypeptide N-Acetylgalactosaminyltransferase 15) has been associated with neuroblastoma proliferation (Hussain, Hoessli, and Fang 2016), and ZNF385A (Zinc Finger Protein 385A) with breast (Walker et al. 2017) and ovarian cancers (Elgaaen et al. 2012).

Of the top 10 genes in the Current/Former analysis,two are known to have links to cancer: QKI (QKI, KH Domain Containing RNA Binding) with tumor suppression(Bandopadhayay et al. 2016) andRPS6KB1 (Ribosomal Protein S6 Kinase B1) withgastrointestinal (Kim et al, 2016) and breast cancer (Sridharan and Basu 2020). Among the top downregulated genes in the Current/Former analysis is LLGL1/HUGL1 (LLGL Scribble Cell Polarity Complex Component 1) which inhibits glioma cell proliferation(Wang et al. 2021). Lower expression (Current/Never analysis) of GALNTL2/GALNT15 (Polypeptide N-Acetylgalactosaminyltransferase 15) has been associated with neuroblastoma proliferation(Hussain, Hoessli, and Fang 2016), and ZNF385A (Zinc Finger Protein 385A) with breast(Walker et al. 2017)and ovarian cancers(Elgaaen et al. 2012).

## Discussion

Weperformed whole transcriptomic analyses to investigate the effects of smoking at the gene expression levelinthe NAcc of postmortem brains in individuals who smoked around the time of death (Current smokers), individuals with a prior history of smoking (Former smokers), and Never smokers. Notable findings include increased expression in neurotransmitter- and cancer-related genes. There were 279 genes that differentiated Current from Former smokers and 105 genes that differentiated Current from Never Smokers after FDR correction. Only two genes differentiated Former from Never smokers suggesting that the effects of smoking on gene expression may be transient.To our knowledge, there is no previously reported human genome-wide gene expression study on the effect of smoking in the NAcc in Current, Former, and Never smokers.

Several recent gene expression studies used post-mortem brain tissue comparing smokers to non-smokers. An interaction between alcohol and smoking was found in the NAcc and VTA of alcoholic smokers, but the main effect of smoking was not clearly determined because of the small number of subjects studied per group and the combination of control subjects and patients with diverse psychiatric diagnoses (Flatscher-Bader et al. 2010, Mexal et al. 2010, Flatscher-Bader et al. 2008, Mexal et al. 2005). In hippocampal tissue of smoking and non-smoking schizophrenics, 277 genes were differentially expressed with many of the largest effect sizes associated with the cholinergic receptor gene family (Mexal et al. 2005). A follow-up qPCR study of the gene CHRNA7 specifically showed an interaction between schizophrenia and smoking (Mexal et al. 2010). To our knowledge, only one study to date investigated gene expression changes in the brain associated with smoking at the time of death in MDDs (Kunii et al. 2015). This studyincluded patients with MDD, BPD, schizophrenia as well as non-affected individuals and evaluated specifically two cholinergic receptor-related genes (CHRFAM7A and CHRNA7) by qPCR. CHRNA7 expression was reduced in schizophrenia and increased in MDD compared to healthy controls with no clear smoking effect on gene expression. Furthermore, none of these studies addressed how smoking interacts with MDD to influencegene expression beyond targeted cholinergic receptor genes. Additionally, to our knowledge, none of the investigations evaluated gene expression differences between former smokers and current (smoking around the time of death) or never smokers.

Many studies have investigated smoking-associated genome wide methylation changes in whole blood (Guida et al. 2015, Joehanes et al. 2016, Ambatipudi et al. 2016, Breitling et al. 2011, Baglietto et al. 2017, Zeilinger et al. 2013), in specific blood cell populations (Su et al. 2016, Reynolds et al. 2017, Sun et al. 2013) and in buccal cells (Teschendorff et al. 2015). These studies produced consistent and reproducible smoking-associated methylation changes particularly in two genes, the aryl-hydrocarbon receptor repressor gene (AHRR) and the F2R like thrombin or trypsin receptor 3 gene (F2RL3), and confirm hypomethylation of these two genes as a smoking biomarker (Gao et al. 2015). Recent studies using an epigenome wide association study (EWAS) approach also confirmed smoking related differential methylation at 14 genes, including AHRR and F2RL3, in blood but also showed their relevance in prediction of cancer risk (Battram et al. 2019). AHRR methylation in blood and saliva can predict smoking status (Current versus Former and Never) and has been proposed as an objective tool to assess smoking status (Philibert et al. 2020, Grieshober et al. 2020). In one comprehensive study, Christensen et al. performed a genome wide methylation analysis in several tissues including brain tissue but in a very small sample of brains (N=12) and blood (N=15) and not from the same subjects making it impossible to perform within subject comparisons of tissue specific alterations associated with smoking (Christensen et al. 2009).A larger EWAS study of five brain regions (anterior cingulate cortex, Brodmann Area 9, caudate nucleus, putamen, and ventral striatum) ranging from 38 to 72 samples per region identified 16 differentially methylated regions and a strong correlation between in matched brain and blood samples in a small (N=10) subsample (Zillich et al. 2022).

The largest number of DEGswere identified when comparing Current to Former smokers.The top Current versus Former genes wereupregulated with no significant differences observed with Never smokers. Many of these genes are metabolic and involved in hypoxia (HIF3A), monocarboxylate (e.g., lactate/beta-hydroxybutyrate) transport (SLC16A1), mitochondrial glutamate transporter (SLC25A18) and Golgi mediated synaptic vesicle fusion (GOSR1). These genes are also potential targets for smoking cessation therapies that could mimic the effects of smoking without the deleterious consequences of cancer related chemicals found in smoke.The upregulation of most genes in our study may be compatible with the upregulating effects of nicotine. It is plausible that a compensatory decrease in glutamatergic activation (and other neurotransmitters) in Former smokers, could account for the wide range of differences in gene expression between the groups. However, more research is needed.

Smoking at the time of death (Current) was associated with an over expression of GABA_A_ family receptor genes: GABRG2 and GABRB3 **(Figure 3)**. Similarly, other GABA related genes such as GAD2, CACNA1E and CACNA2D3 were also upregulated in Current smokers versus Never smokers. Similar changes were also observed in the glutamate transporter gene (SLC1A1) and the AMPA type glutamate receptor GRIA2. Interestingly, expression of another intracellular glutamate transporter gene (SLC25A18), involved in the transport of glutamate across the inner mitochondrial membrane, was also altered in Current smokers versus Former smokers. However, no significant change was observed compared to Never smokers, suggesting this glutamate transporter is normally downregulated in subjects susceptible to smoking dependence (Former) but who no longer smoke. Low expression of the SLC25A18 glutamate transporter might, therefore, be a marker of smoking addiction susceptibility **(Figure 3)**. These results suggest that the increased depolarization and dopamine release in the NAcc of smokers results in possible compensatory mechanisms involving an overall increase of GABA inhibitory neurotransmission. The observed increase in GRIA2 expression and glutamate transporter also suggests compensatory effects of increased tobacco related over-activation of the NAcc. Finally, the dopaminergic receptor genes (DRD1 and DRD2) and cholinergic receptor genes (CHRNA3 and CHRNB2) were found to be reliably detected in the NAcc in our data, however only the CHRNB2 receptor was found to bedifferentially expressed. CHRNB2 receptor expression was significantly decreased among Current smokers when compared to both Former and Never smokers (**Figure 3**). The relatively few genes that differentiated Former and Never smokers suggest transcriptomic alterations may revert to control levels following smoking cessation. Our findings are consistent with observations in both brain and blood comparing current and former smokers (Joehanes et al. 2016). Some non-transcriptomic comparisons of former and never smokers support the hypothesis of quick reversion to the non-smoker physiological state. SPECT imaging of nicotinic receptor binding levels in current smokers show a reduction to non-smoker levels within 21 days of smoking cessation(Mamede et al. 2007), and binding in post-mortem hippocampus and thalamus from subjects who quit smoking, return to non-smoker levels within 2 months (Breese et al. 1997). In contrast, lasting alterations in the physiology of addiction pathways have been found in the cAMP signal transduction of the post-mortem VTA of both current and former smokers compared to non-smokers (Hope et al. 2007). Longer-term effects were found with microstructural white matter changes in cerebral white matter with diffusion tensor imaging parameters not returning to non-smoker levels until at least 20 years after cessation (Gons et al. 2011).

The IPA analysis revealed an unexpectedly large number of cancer related genes in comparisons between Current smokers and both non-smoking groups (Current versus Former and Current versus Never). This is particularly interesting because the subjects included in this study were carefully reviewed to exclude cancer using medical records, pathological observations, and a neuropathological review of the brain. Thus, cancer-related genes seem to be activated specifically in Current smokers but not in non-smoker groups. Former smokers lack of cancer related signature when compared to Never smokers suggestssmoking cessation can lead to the reversal of an oncogenic signature observed in Current smokers.

Several functional studies have previously explored gene expression(Flatscher-Bader et al. 2010, Mexal et al. 2010, Flatscher-Bader et al. 2008, Mexal et al. 2005) and smoking-associated genome wide methylation changes in whole blood (Guida et al. 2015, Joehanes et al. 2016, Ambatipudi et al. 2016, Breitling et al. 2011, Baglietto et al. 2017, Zeilinger et al. 2013), in specific blood cell populations (Su et al. 2016, Reynolds et al. 2017, Sun et al. 2013) and in buccal cells (Teschendorff et al. 2015). When comparing significant differentially expressed genes in the NAcc to previously identified smoking-associated genes, we observe considerable overlap **(Supplementary Figure 4)** with previous studies.

One methylation study by Markunas et al. (Markunas et al. 2021) used NAcc and directly compared smokers to non-smokers. Of the seven genes identified by Markunas and colleagues(Markunas et al. 2021), our data showed an overlap with five of the seven genes (ZCCHC24, CTCF, MTMR6, PRKD) suggesting methylation changes caused by smoking can lead to significant corresponding changes at the mRNA level. Mexal et al. (Mexal et al. 2005)performed a similar study to ours by comparing genome vide expression of smokers and non-smokers in the hippocampus using an older version of Affymetrix microarrays (Hu95Av2 chips, containing 12,625), we used data from the Affymetrix HG-U133 Plus2.0 chips. Mexal et al also included some subjects in smoker and non-smoker groups that had cancer(Mexal et al. 2005), while we excluded any subjects with any evidence of cancer by examining medical files and neuropathological examination of the brain. While Mexal et al.(Mexal et al. 2005) and our study differ in terms of the microarray chips used and the brain region investigated (hippocampus versus NAcc), we observed an important overlap of genes differentially expressed between the two studies. In Mexal et al.(Mexal et al. 2005)the authors reported 277 differentially expressed genes in the hippocampus of smokers versus non-smokers. After re-annotation of these transcripts (using Genecards and Omim) we identified 257 currently annotated genes with gene symbols of which 83 unique genes that overlap between the two studies (32%). Furthermore, both in Mexal et al.(Mexal et al. 2005) and in our study several GABAergic and glutamatergic genes (GABRB3, GRIA1) as well as some of the cancer related genes are found to be increased in smokers suggesting a similar up-regulation of these pathways because of smoking.

Psychiatric patients were included, in addition to controls, as this population has a high incidence of nicotine dependence(Fornaro et al. 2022). Whilethe small sample size precluded the investigation of within-diagnostic effects of smoking, diagnoses were controlled for in theanalyses.We recognized significant differences in age (Current smokers were younger) and gender (greater number of females in the Never smoking group) as potential confoundsand controlled for them in our analyses.Also, the high co-morbidity between smoking and alcohol could be a factor in the results although alcohol abuse was ruled out by our psychological autopsy. Further analyses in this same study suggested that age, pH, PMI and possibly gender did not influence the findings in the NAcc. Analyses of inclusion or exclusion of females (10%) by othersdid not alter theirmicroarray results(Flatscher-Bader et al. 2010).

Further research will be needed to address some of the limitations of this study. First, while mood disorders have been associated with smoking in many studies, we focused on overall smoking effects without addressing diagnostic specific effects due to the small sample size. Second, a larger sample will be needed in future studies to explore possible gender differences and investigate potentialmedication effects. Finally, in addition to the NAcc, other brain regions involved in addictionsuch as amygdala, ventral tegmental area and prefrontal areas, stress, and impulse regulation, should also be investigated to gain a more comprehensive understanding of the circuit dysregulations associated with smoking in the human brain.

## Data Availability

All data produced in the present study are available upon reasonable request to the authors.

## Acknowledgements

The authors are members of the Pritzker Neuropsychiatric Disorders Research Consortium, which is supported by the Pritzker Neuropsychiatric Disorders Research Fund. A shared intellectual property agreement exists between this philanthropic fund and the University of Michigan, Stanford University, the Weill Medical College of Cornell University, HudsonAlpha Institute of Biotechnology, and the University of California Irvine, to encourage the development of appropriate findings for research and clinical applications. We are also grateful to David Walsh and Casey Kathleen Burke for characterizing, acquiring, and storing the postmortem human tissue.

## Author Contributions

AS, RS, MPV, and WEB conceived and designed the experiments. BB, PC, RS, and WEB were involved in collection of post-mortem brain and blood, and performance of psychological autopsies. AS analyzed the data. RS, FM, BB, MPV, WEB, FSL, JB, AFS, RM, SJW, and HA contributed to manuscript preparation, reviewing, and editing. All authors reviewed and approved the final manuscript.

## Data Availability Statement

All data produced in the present study are available upon reasonable request to the authors

## Supplementary Material for

**Supplementary Table 1:** Demographics and post-mortem interval (PMI)

| Smoking Group | N | Male | Female | Age (Mean(SD)) | PMI (Mean(SD)) |
| --- | --- | --- | --- | --- | --- |
| Current | 20 | 18 | 2 | 44.2(13.5)* | 22.4(8.4) |
| Former | 13 | 12 | 1 | 55.5(16.5) | 21.8(9.5) |
| Never | 18 | 11 | 7^ | 52.3(11.8) | 20.8(5.9) |
| **Total** | **51** | **41** | **10** | **50.0(14.3)** | **21.7(7.8)** |

\*Current smokers younger than Former (p=0.032) and Never (p=0.048)
^Higher percentage of females among Never smokers (X=7.14, p=0.028)

**Supplementary Table 2:** Sample diagnostic and demographic details.

| Smoking Status | Count | BP | Non-psychiatric | MDD |
| --- | --- | --- | --- | --- |
| Never | 18 | 4 | 10 | 4 |
| Former | 13 | 1 | 8 | 4 |
| Current | 20 | 3 | 4 | 13 |
| **TOTAL** | **51** | **8** | **22** | **21** |

**Supplementary Table 3.** Number of DEGs (q-value≤ 0.05) and direction of change by smoking status.

|  | Total | # UP | # DOWN |
| --- | --- | --- | --- |
| Current to Former | 279 | 265 | 14 |
| Current to Never | 105 | 101 | 4 |
| Former to Never | 2 | 0 | 2 |

**Supplementary Table 4:**
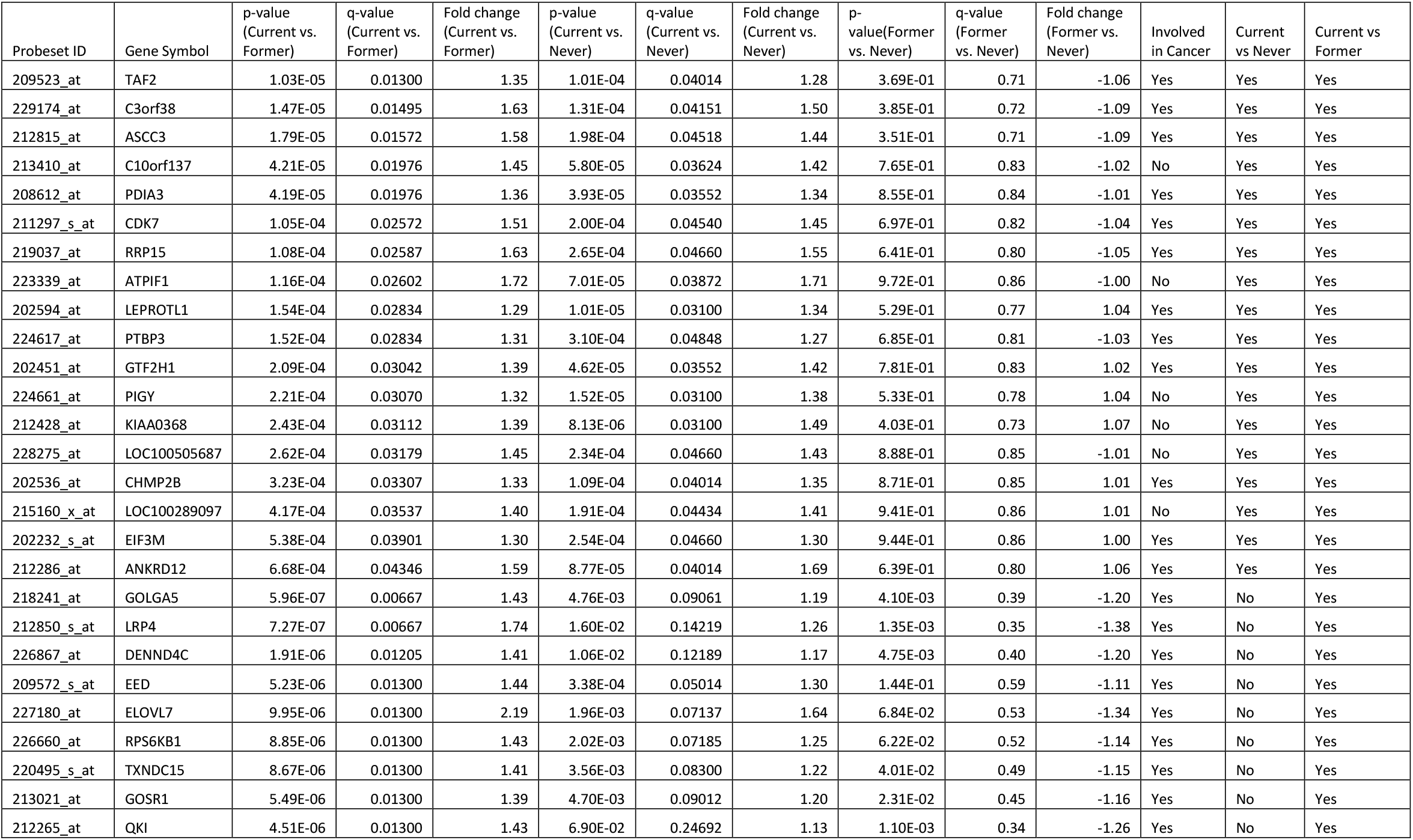
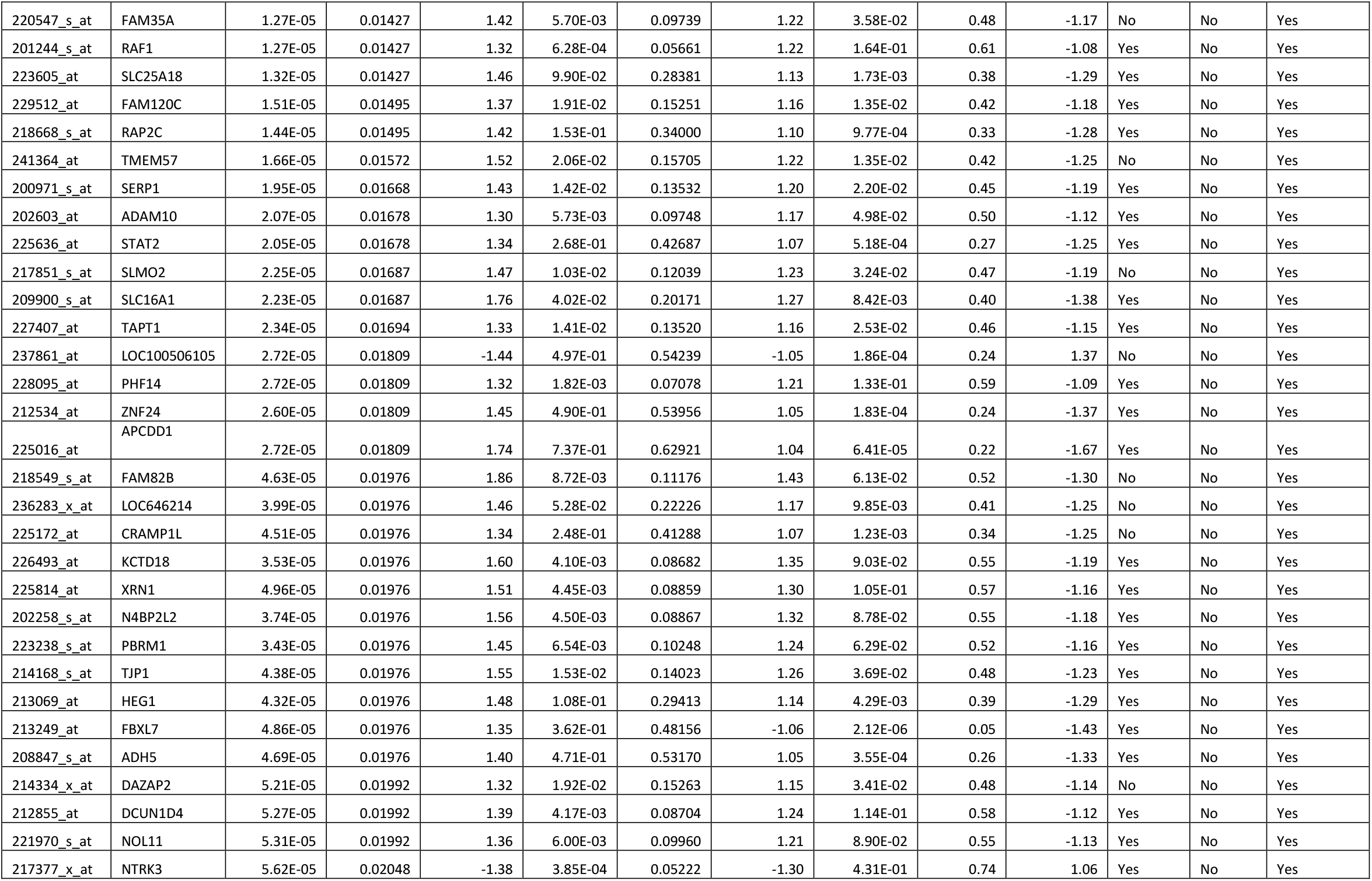
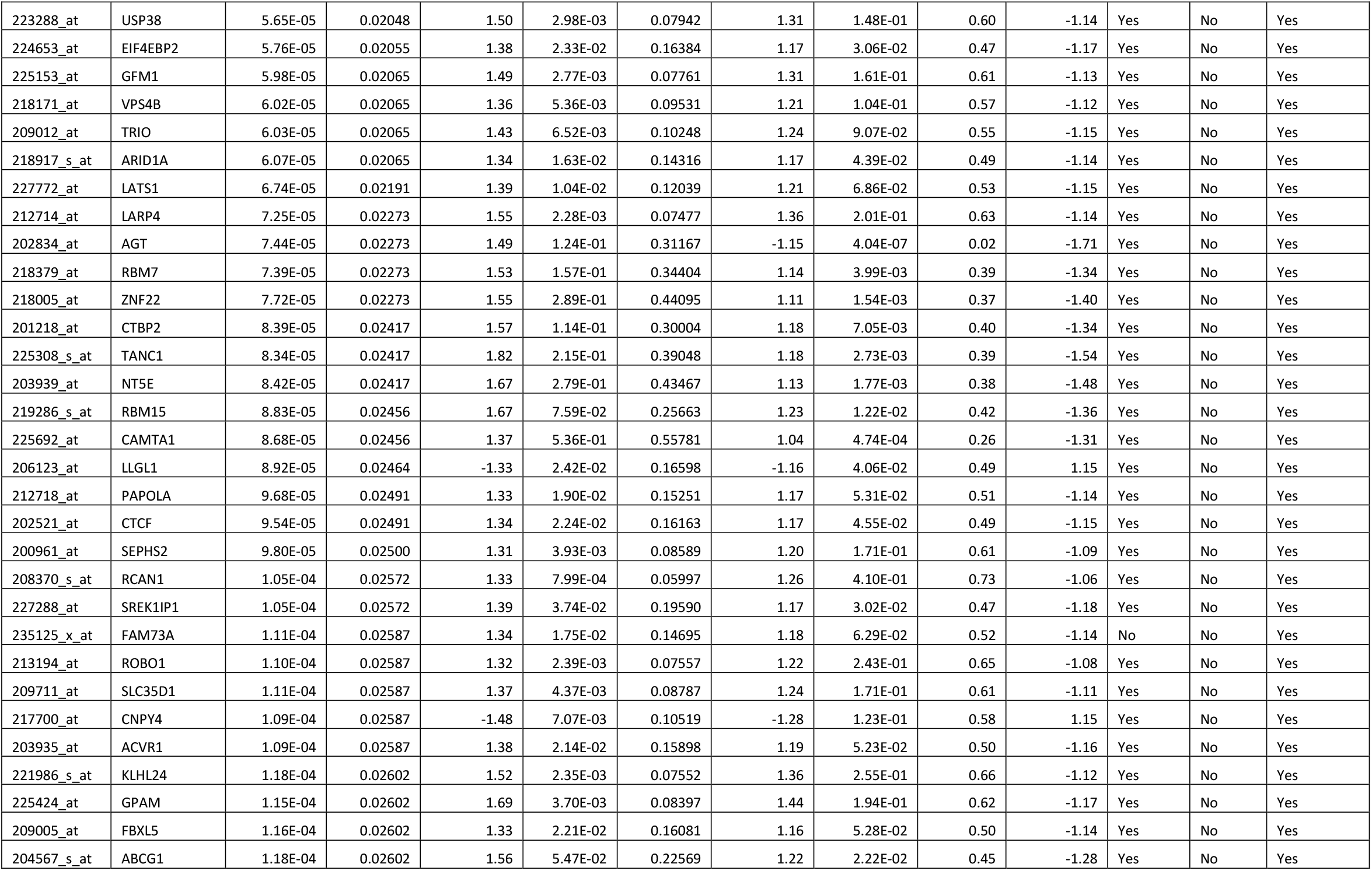
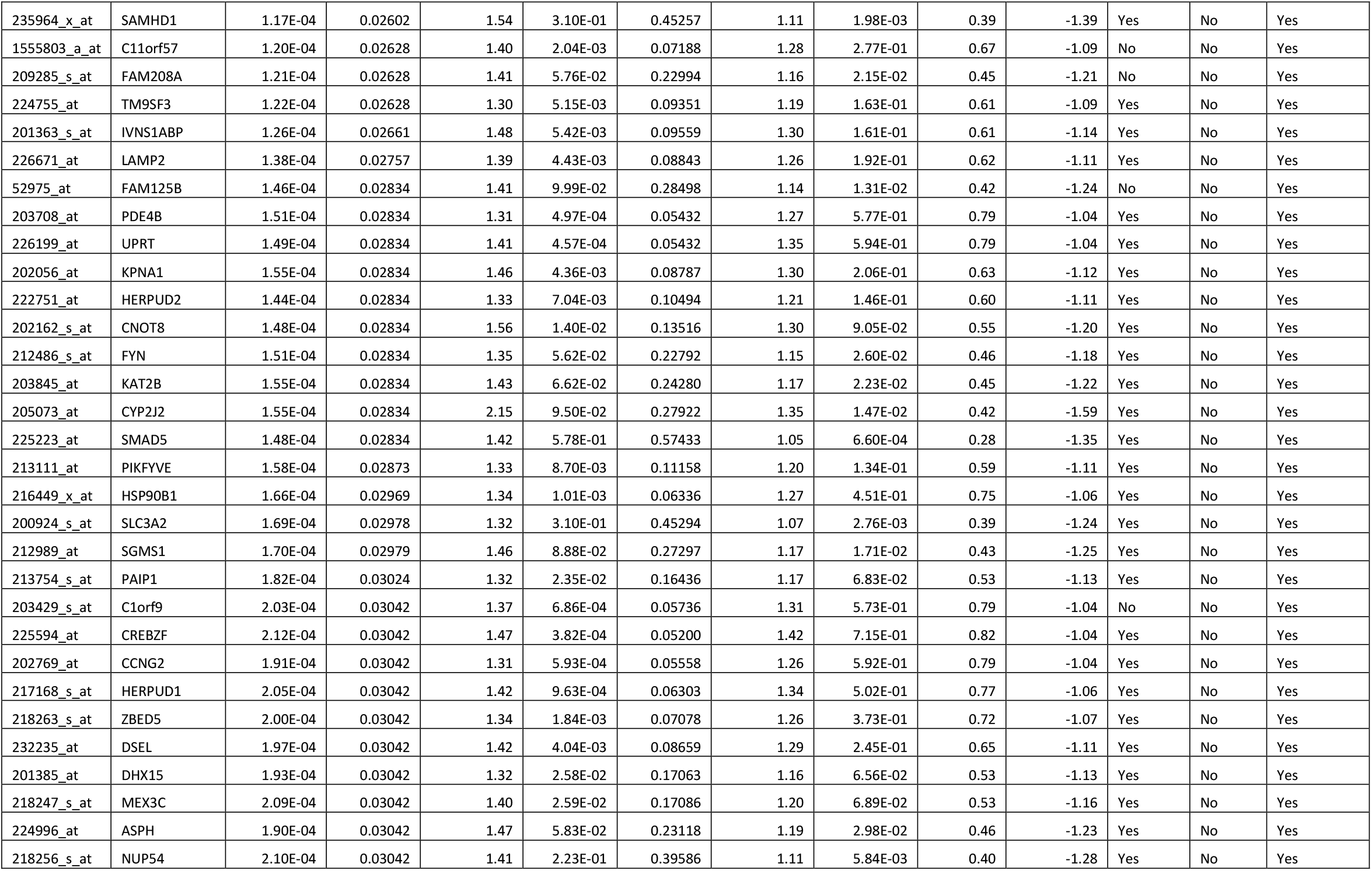
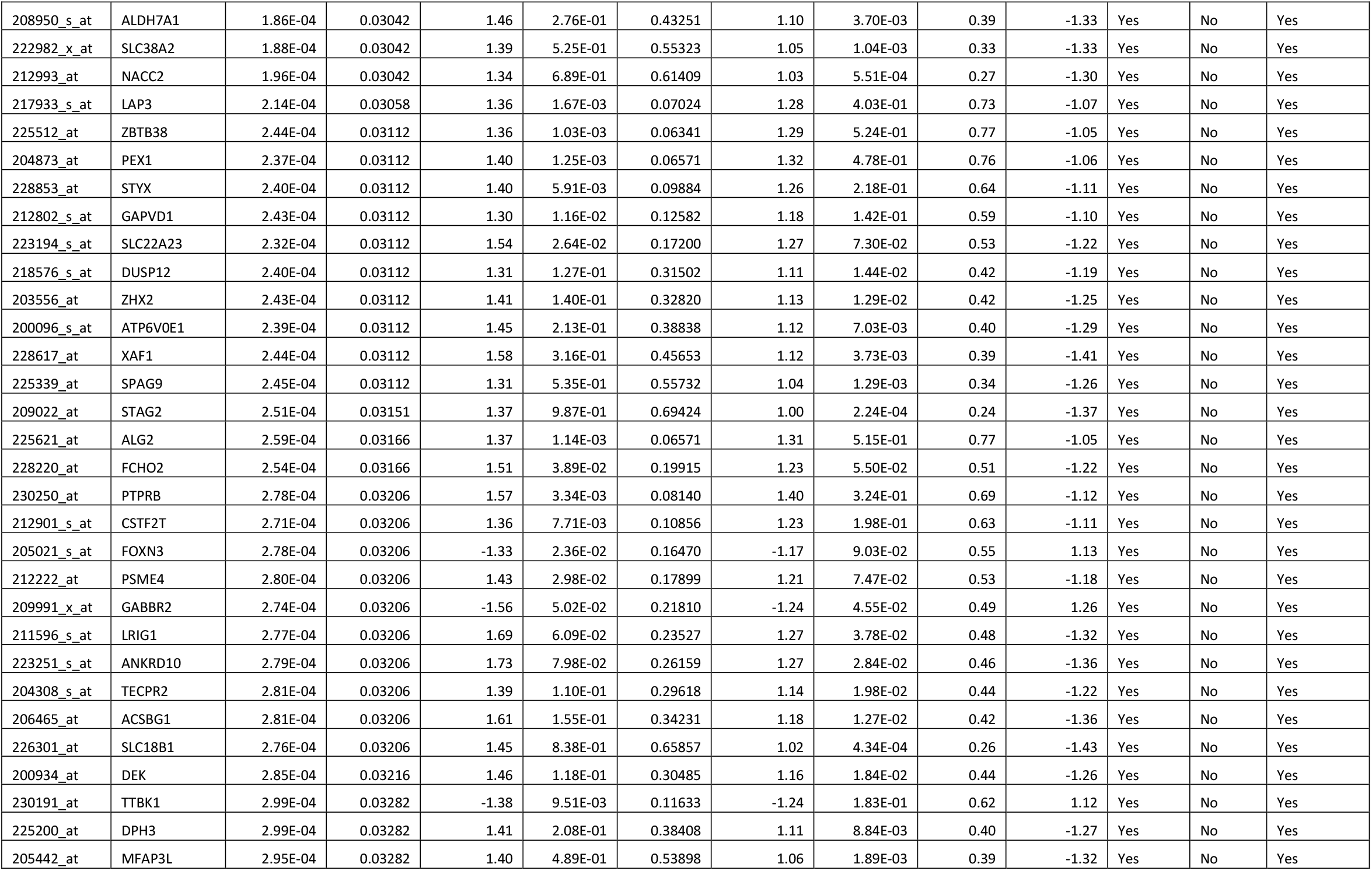
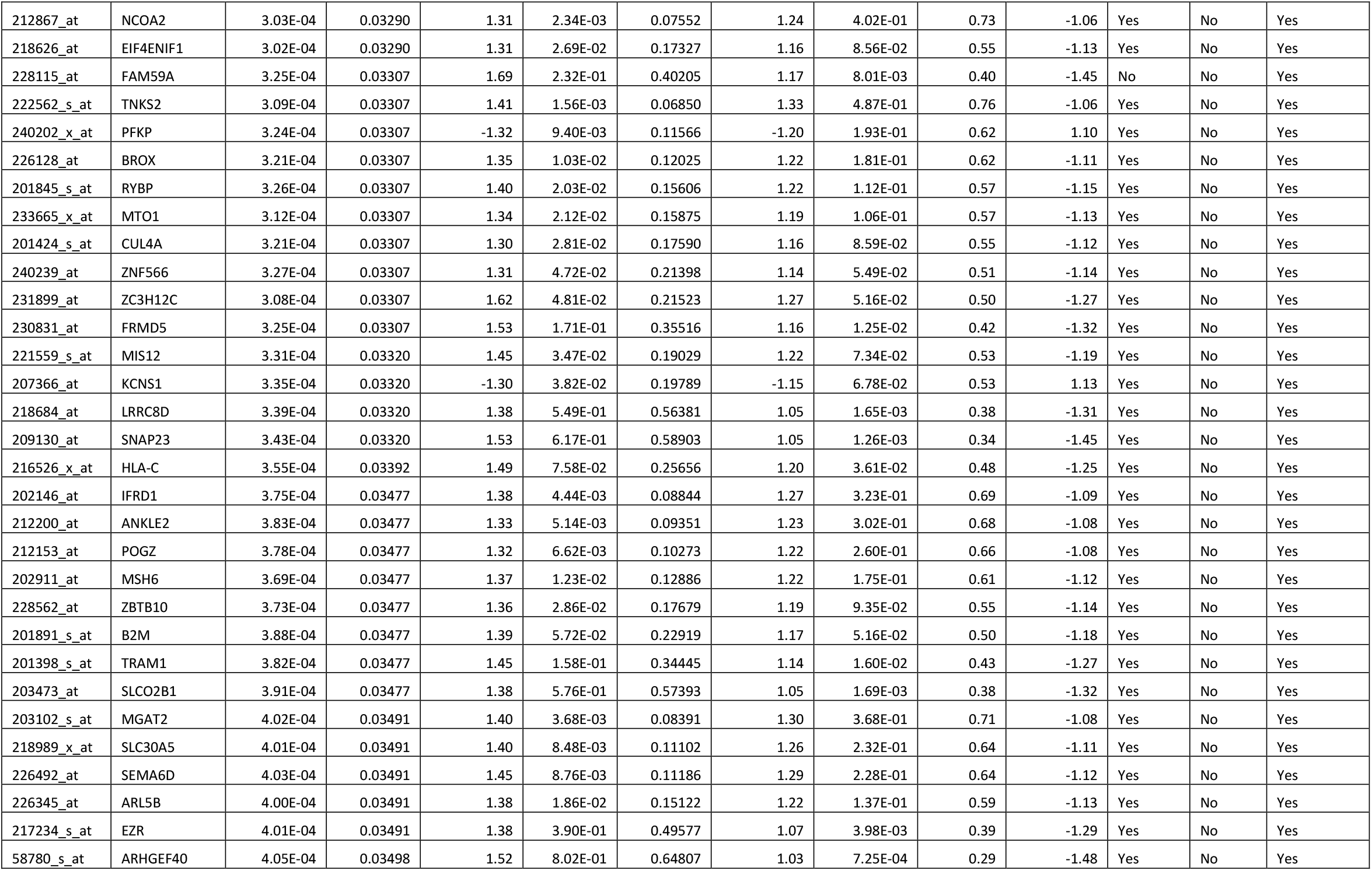
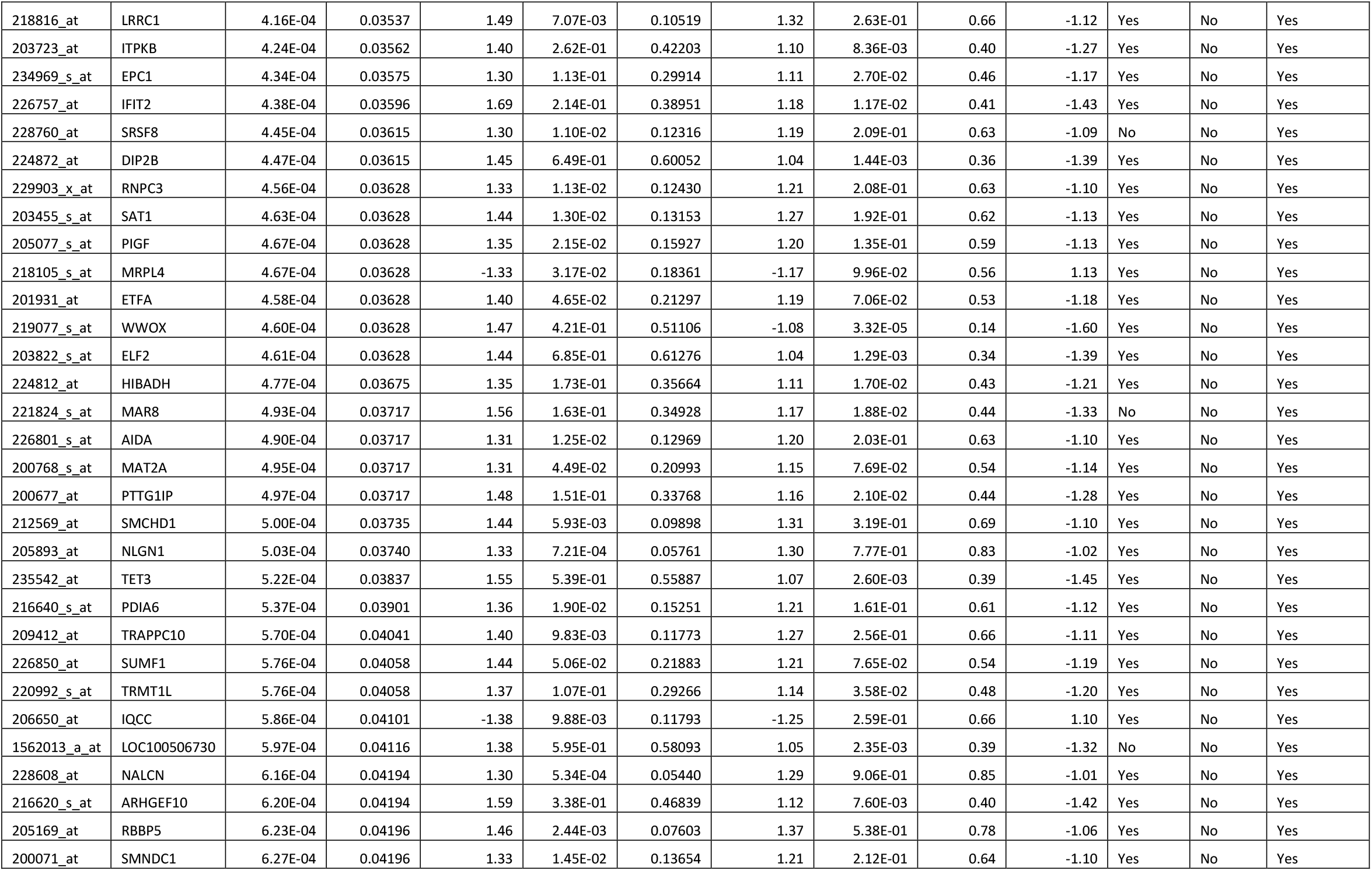
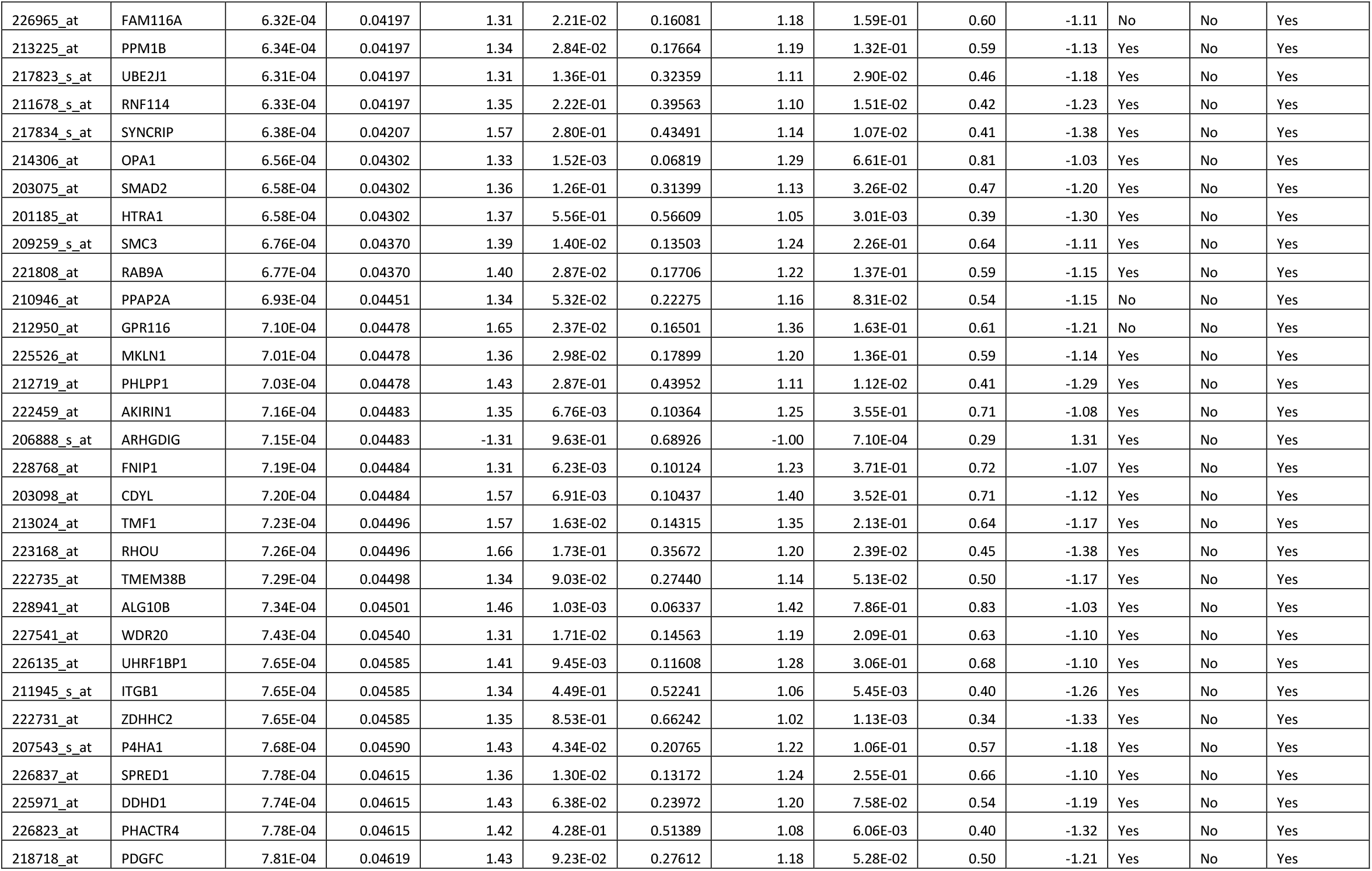
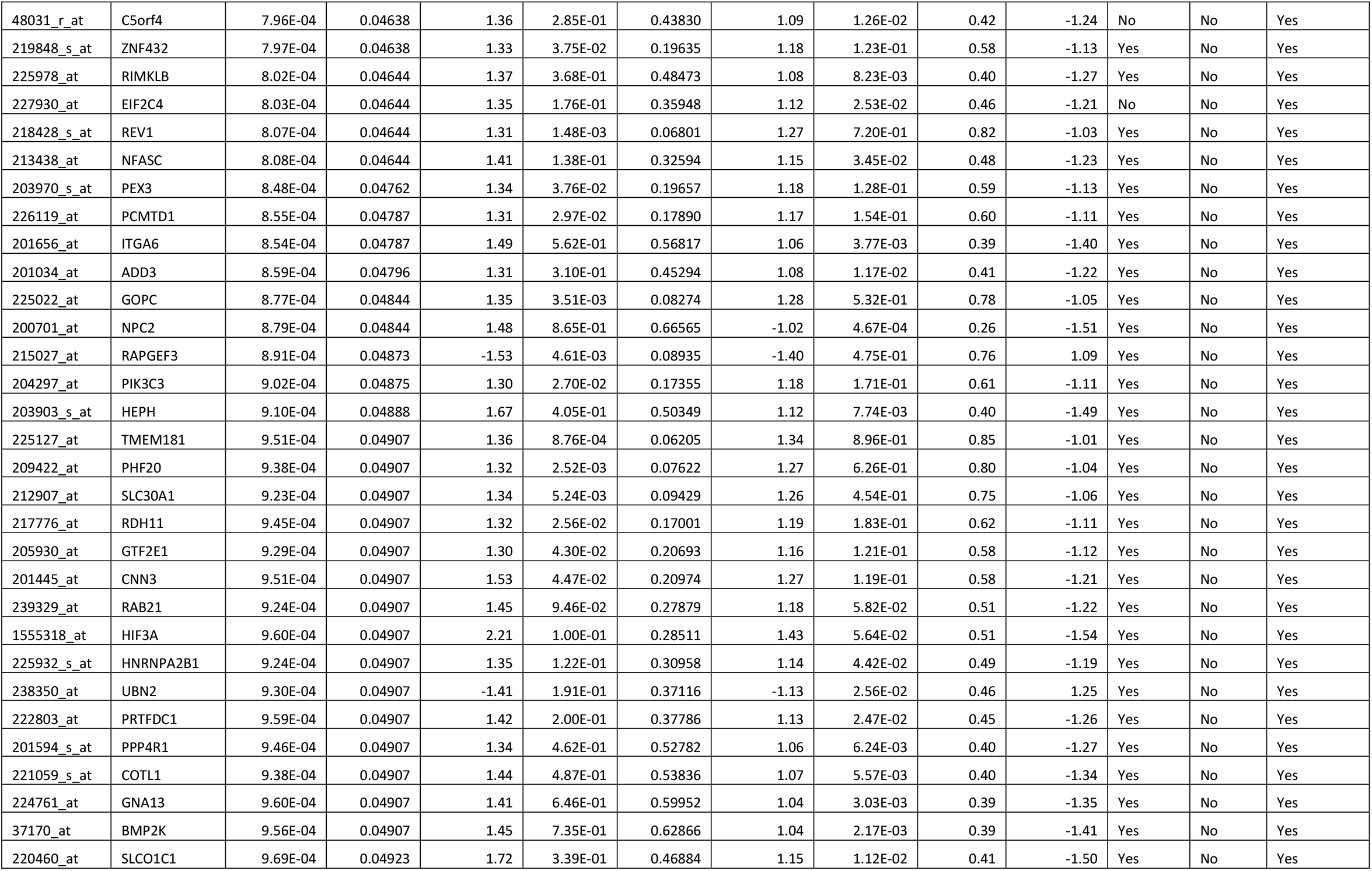
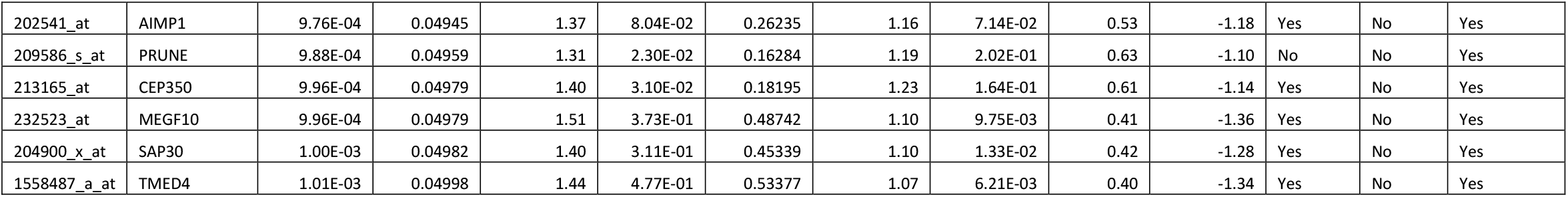
Differentially expressed probesets (genes) between Current vs Former smokers. P-values, corresponding FDR corrected q-values (<0.05) and fold changes are presented by comparison. Involvement in Cancer/Organismal Injury and abnormalities is also presented.

**Supplementary Table 5:** Differentially expressed probesets (genes) between Current vs Never smokers. P-values, corresponding FDR corrected q-values and fold changes are presented by comparison. Involvement in Cancer/Organismal Injury and abnormalities is also presented.

| Probeset ID | Gene Symbol | p-value<br>(Current vs.<br>Former) | q-value<br>(Current<br>vs. Former) | Fold change<br>(Current vs.<br>Former) | p-value<br>(Current<br>vs. Never) | q-value<br>(Current<br>vs. Never) | Fold change<br>(Current vs.<br>Never) | p-value<br>(Former<br>vs. Never) | q-value<br>(Former<br>vs. Never) | Fold change<br>(Former vs.<br>Never) | Involved<br>in Cancer | Current<br>vs Never | Current vs<br>Former |
| --- | --- | --- | --- | --- | --- | --- | --- | --- | --- | --- | --- | --- | --- |
| 202214_s_at | CUL4B | 4.20E-03 | 0.07908 | 1.19 | 2.78E-06 | 0.03095 | 1.33 | 3.77E-02 | 0.48 | 1.13 | Yes | Yes | No |
| 209409_at | GRB10 | 4.26E-03 | 0.07947 | 1.17 | 1.92E-06 | 0.03095 | 1.31 | 2.90E-02 | 0.46 | 1.12 | Yes | Yes | No |
| 219297_at | WDR44 | 9.39E-03 | 0.10670 | 1.24 | 5.13E-06 | 0.03095 | 1.49 | 2.79E-02 | 0.46 | 1.20 | Yes | Yes | No |
| 224661_at | PIGY | 2.21E-04 | 0.03070 | 1.32 | 1.52E-05 | 0.03100 | 1.38 | 5.33E-01 | 0.78 | 1.04 | No | Yes | Yes |
| 212428_at | KIAA0368 | 2.43E-04 | 0.03112 | 1.39 | 8.13E-06 | 0.03100 | 1.49 | 4.03E-01 | 0.73 | 1.07 | No | Yes | Yes |
| 202594_at | LEPROTL1 | 1.54E-04 | 0.02834 | 1.29 | 1.01E-05 | 0.03100 | 1.34 | 5.29E-01 | 0.77 | 1.04 | Yes | Yes | Yes |
| 225391_at | LOC93622 | 6.65E-03 | 0.09270 | 1.23 | 6.93E-06 | 0.03100 | 1.42 | 4.62E-02 | 0.49 | 1.16 | No | Yes | No |
| 227690_at | GABRB3 | 3.60E-02 | 0.17922 | 1.16 | 1.15E-05 | 0.03100 | 1.38 | 1.27E-02 | 0.42 | 1.19 | Yes | Yes | No |
| 218764_at | PRKCH | 4.20E-02 | 0.18953 | 1.27 | 1.42E-05 | 0.03100 | 1.71 | 1.25E-02 | 0.42 | 1.34 | Yes | Yes | No |
| 229300_at | RAB3C | 5.41E-02 | 0.21149 | 1.20 | 1.55E-05 | 0.03100 | 1.52 | 9.87E-03 | 0.41 | 1.27 | Yes | Yes | No |
| 235077_at | MEG3 | 9.57E-02 | 0.26411 | 1.18 | 1.28E-05 | 0.03100 | 1.58 | 4.06E-03 | 0.39 | 1.34 | Yes | Yes | No |
| 208612_at | PDIA3 | 4.19E-05 | 0.01976 | 1.36 | 3.93E-05 | 0.03552 | 1.34 | 8.55E-01 | 0.84 | -1.01 | Yes | Yes | Yes |
| 202451_at | GTF2H1 | 2.09E-04 | 0.03042 | 1.39 | 4.62E-05 | 0.03552 | 1.42 | 7.81E-01 | 0.83 | 1.02 | Yes | Yes | Yes |
| 208798_x_at | GOLGA8A | 1.92E-03 | 0.05984 | 1.29 | 2.02E-05 | 0.03552 | 1.43 | 2.07E-01 | 0.63 | 1.10 | No | Yes | No |
| 200629_at | WARS | 1.22E-02 | 0.11770 | 1.23 | 5.16E-05 | 0.03552 | 1.41 | 9.72E-02 | 0.56 | 1.14 | No | Yes | No |
| 203721_s_at | UTP18 | 1.09E-03 | 0.05135 | 1.29 | 5.31E-05 | 0.03552 | 1.37 | 4.33E-01 | 0.74 | 1.06 | Yes | Yes | No |
| 202724_s_at | FOXO1 | 1.19E-03 | 0.05282 | 1.28 | 2.50E-05 | 0.03552 | 1.38 | 3.01E-01 | 0.68 | 1.08 | Yes | Yes | No |
| 227525_at | GLCCI1 | 1.59E-03 | 0.05715 | 1.30 | 2.77E-05 | 0.03552 | 1.42 | 2.69E-01 | 0.67 | 1.09 | Yes | Yes | No |
| 226111_s_at | ZNF385A | 2.50E-03 | 0.06611 | -1.23 | 5.20E-05 | 0.03552 | -1.31 | 2.81E-01 | 0.67 | -1.07 | Yes | Yes | No |
| 221918_at | CDK17 | 4.32E-03 | 0.07977 | 1.21 | 4.10E-05 | 0.03552 | 1.31 | 1.80E-01 | 0.62 | 1.09 | Yes | Yes | No |
| 225996_at | LONRF2 | 5.21E-03 | 0.08581 | 1.23 | 4.60E-05 | 0.03552 | 1.35 | 1.69E-01 | 0.61 | 1.10 | Yes | Yes | No |
| 216088_s_at | PSMA7 | 9.32E-03 | 0.10628 | 1.20 | 4.97E-05 | 0.03552 | 1.34 | 1.17E-01 | 0.58 | 1.11 | Yes | Yes | No |
| 206052_s_at | SLBP | 1.77E-02 | 0.13620 | 1.17 | 2.49E-05 | 0.03552 | 1.34 | 4.49E-02 | 0.49 | 1.14 | Yes | Yes | No |
| 202254_at | SIPA1L1 | 2.56E-02 | 0.15727 | 1.17 | 3.92E-05 | 0.03552 | 1.34 | 4.29E-02 | 0.49 | 1.15 | Yes | Yes | No |
| 228824_s_at | PTGR1 | 1.21E-01 | 0.29002 | 1.22 | 4.58E-05 | 0.03552 | 1.71 | 8.06E-03 | 0.40 | 1.41 | Yes | Yes | No |
| 202334_s_at | UBE2B | 5.45E-03 | 0.08652 | 1.21 | 3.82E-05 | 0.03552 | 1.32 | 1.48E-01 | 0.60 | 1.10 | Yes | Yes | No |
| 213410_at | C10orf137 | 4.21E-05 | 0.01976 | 1.45 | 5.80E-05 | 0.03624 | 1.42 | 7.65E-01 | 0.83 | -1.02 | No | Yes | Yes |
| 223606_x_at | KIAA1704 | 2.98E-03 | 0.07017 | 1.27 | 6.28E-05 | 0.03686 | 1.38 | 2.78E-01 | 0.67 | 1.09 | No | Yes | No |
| 213411_at | ADAM22 | 2.35E-02 | 0.15223 | 1.21 | 6.03E-05 | 0.03686 | 1.41 | 6.16E-02 | 0.52 | 1.17 | Yes | Yes | No |
| 228423_at | MAP9 | 1.31E-03 | 0.05407 | 1.26 | 6.45E-05 | 0.03686 | 1.32 | 4.29E-01 | 0.74 | 1.05 | Yes | Yes | No |
| 223339_at | ATPIF1 | 1.16E-04 | 0.02602 | 1.72 | 7.01E-05 | 0.03872 | 1.71 | 9.72E-01 | 0.86 | -1.00 | No | Yes | Yes |
| 213664_at | SLC1A1 | 4.69E-02 | 0.19864 | 1.16 | 7.24E-05 | 0.03941 | 1.36 | 3.55E-02 | 0.48 | 1.17 | Yes | Yes | No |
| 229309_at | ADRB1 | 2.48E-02 | 0.15544 | 1.24 | 7.65E-05 | 0.03992 | 1.47 | 6.82E-02 | 0.53 | 1.19 | Yes | Yes | No |
| 209523_at | TAF2 | 1.03E-05 | 0.01300 | 1.35 | 1.01E-04 | 0.04014 | 1.28 | 3.69E-01 | 0.71 | -1.06 | Yes | Yes | Yes |
| 202536_at | CHMP2B | 3.23E-04 | 0.03307 | 1.33 | 1.09E-04 | 0.04014 | 1.35 | 8.71E-01 | 0.85 | 1.01 | Yes | Yes | Yes |
| 212286_at | ANKRD12 | 6.68E-04 | 0.04346 | 1.59 | 8.77E-05 | 0.04014 | 1.69 | 6.39E-01 | 0.80 | 1.06 | Yes | Yes | Yes |
| 218499_at | MST4 | 1.22E-01 | 0.29077 | 1.21 | 1.07E-04 | 0.04014 | 1.64 | 1.54E-02 | 0.42 | 1.36 | No | Yes | No |
| 201634_s_at | CYB5B | 1.61E-03 | 0.05740 | 1.29 | 1.03E-04 | 0.04014 | 1.36 | 4.70E-01 | 0.75 | 1.06 | Yes | Yes | No |
| 225485_at | CEP41 | 6.43E-03 | 0.09178 | 1.26 | 1.04E-04 | 0.04014 | 1.39 | 2.25E-01 | 0.64 | 1.10 | Yes | Yes | No |
| 202049_s_at | ZMYM4 | 6.43E-03 | 0.09181 | 1.28 | 9.79E-05 | 0.04014 | 1.43 | 2.18E-01 | 0.64 | 1.11 | Yes | Yes | No |
| 227871_at | CHM | 2.72E-02 | 0.16055 | 1.18 | 1.03E-04 | 0.04014 | 1.33 | 7.56E-02 | 0.53 | 1.14 | Yes | Yes | No |
| 226888_at | CSNK1G1 | 4.88E-02 | 0.20182 | 1.27 | 1.09E-04 | 0.04014 | 1.62 | 4.49E-02 | 0.49 | 1.27 | Yes | Yes | No |
| 238601_at | PHKB | 5.22E-02 | 0.20800 | 1.28 | 1.04E-04 | 0.04014 | 1.65 | 4.08E-02 | 0.49 | 1.29 | Yes | Yes | No |
| 203139_at | DAPK1 | 6.01E-02 | 0.22013 | 1.17 | 1.07E-04 | 0.04014 | 1.39 | 3.56E-02 | 0.48 | 1.19 | Yes | Yes | No |
| 229129_at | HNRNPD | 8.66E-02 | 0.25348 | 1.14 | 9.91E-05 | 0.04014 | 1.36 | 2.23E-02 | 0.45 | 1.19 | Yes | Yes | No |
| 1552767_a_at | HS6ST2 | 1.19E-01 | 0.28841 | 1.18 | 1.04E-04 | 0.04014 | 1.53 | 1.54E-02 | 0.42 | 1.30 | Yes | Yes | No |
| 213913_s_at | TBC1D30 | 1.40E-01 | 0.30841 | 1.16 | 1.02E-04 | 0.04014 | 1.49 | 1.23E-02 | 0.42 | 1.29 | Yes | Yes | No |
| 219714_s_at | CACNA2D3 | 1.80E-01 | 0.34114 | 1.18 | 1.04E-04 | 0.04014 | 1.65 | 8.67E-03 | 0.40 | 1.40 | Yes | Yes | No |
| 208638_at | PDIA6 | 2.92E-03 | 0.06965 | 1.23 | 8.44E-05 | 0.04014 | 1.31 | 3.22E-01 | 0.69 | 1.07 | Yes | Yes | No |
| 239461_at | GALNTL2 | 8.10E-01 | 0.62444 | -1.02 | 1.12E-04 | 0.04034 | -1.50 | 3.64E-04 | 0.26 | -1.46 | No | Yes | No |
| 222393_s_at | NAA50 | 4.20E-02 | 0.18967 | 1.23 | 1.14E-04 | 0.04034 | 1.48 | 5.40E-02 | 0.51 | 1.21 | Yes | Yes | No |
| 217949_s_at | VKORC1 | 2.07E-03 | 0.06192 | 1.26 | 1.20E-04 | 0.04145 | 1.33 | 4.45E-01 | 0.75 | 1.06 | Yes | Yes | No |
| 229174_at | C3orf38 | 1.47E-05 | 0.01495 | 1.63 | 1.31E-04 | 0.04151 | 1.50 | 3.85E-01 | 0.72 | -1.09 | Yes | Yes | Yes |
| 218259_at | MKL2 | 6.83E-02 | 0.23129 | 1.15 | 1.36E-04 | 0.04151 | 1.34 | 3.66E-02 | 0.48 | 1.17 | No | Yes | No |
| 212887_at | SEC23A | 2.95E-03 | 0.06987 | 1.25 | 1.36E-04 | 0.04151 | 1.33 | 3.96E-01 | 0.73 | 1.06 | Yes | Yes | No |
| 212990_at | SYNJ1 | 2.69E-02 | 0.15998 | 1.17 | 1.28E-04 | 0.04151 | 1.31 | 8.75E-02 | 0.55 | 1.12 | Yes | Yes | No |
| 204072_s_at | FRY | 8.20E-02 | 0.24799 | 1.12 | 1.27E-04 | 0.04151 | 1.30 | 2.84E-02 | 0.46 | 1.16 | Yes | Yes | No |
| 228284_at | TLE1 | 8.58E-02 | 0.25240 | 1.26 | 1.26E-04 | 0.04151 | 1.71 | 2.67E-02 | 0.46 | 1.35 | Yes | Yes | No |
| 225930_at | NKIRAS1 | 3.58E-01 | 0.45377 | 1.08 | 1.26E-04 | 0.04151 | 1.38 | 3.09E-03 | 0.39 | 1.28 | Yes | Yes | No |
| 221874_at | KIAA1324 | 2.12E-02 | 0.14718 | 1.32 | 1.49E-04 | 0.04242 | 1.59 | 1.17E-01 | 0.58 | 1.20 | No | Yes | No |
| 219685_at | TMEM35 | 2.09E-01 | 0.36402 | 1.13 | 1.48E-04 | 0.04242 | 1.47 | 9.05E-03 | 0.40 | 1.30 | No | Yes | No |
| 223283_s_at | TSHZ1 | 1.76E-03 | 0.05887 | 1.41 | 1.54E-04 | 0.04291 | 1.50 | 5.28E-01 | 0.77 | 1.07 | Yes | Yes | No |
| 202557_at | HSPA13 | 9.46E-03 | 0.10693 | 1.22 | 1.53E-04 | 0.04291 | 1.34 | 2.13E-01 | 0.64 | 1.10 | Yes | Yes | No |
| 203884_s_at | RAB11FIP2 | 7.82E-03 | 0.09925 | 1.21 | 1.58E-04 | 0.04320 | 1.30 | 2.45E-01 | 0.65 | 1.08 | Yes | Yes | No |
| 215160_x_at | LOC100289097 | 4.17E-04 | 0.03537 | 1.40 | 1.91E-04 | 0.04434 | 1.41 | 9.41E-01 | 0.86 | 1.01 | No | Yes | Yes |
| 229328_at | LOC100507433 | 1.20E-03 | 0.05290 | 1.30 | 1.92E-04 | 0.04434 | 1.34 | 6.67E-01 | 0.81 | 1.03 | No | Yes | No |
| 203816_at | DGUOK | 8.80E-03 | 0.10366 | 1.32 | 1.92E-04 | 0.04434 | 1.48 | 2.51E-01 | 0.65 | 1.12 | Yes | Yes | No |
| 235604_x_at | ZNF493 | 2.79E-02 | 0.16254 | 1.18 | 1.68E-04 | 0.04434 | 1.32 | 1.00E-01 | 0.56 | 1.13 | Yes | Yes | No |
| 228775_at | EMC3 | 7.24E-02 | 0.23609 | 1.21 | 1.78E-04 | 0.04434 | 1.50 | 4.15E-02 | 0.49 | 1.24 | Yes | Yes | No |
| 213547_at | CAND2 | 7.47E-02 | 0.23860 | 1.14 | 1.89E-04 | 0.04434 | 1.33 | 4.18E-02 | 0.49 | 1.16 | Yes | Yes | No |
| 238747_at | CACNA1E | 3.00E-01 | 0.42139 | 1.11 | 1.64E-04 | 0.04434 | 1.47 | 5.39E-03 | 0.40 | 1.33 | Yes | Yes | No |
| 1555882_at | SPIN3 | 4.56E-01 | 0.50004 | 1.17 | 1.89E-04 | 0.04434 | 2.21 | 2.66E-03 | 0.39 | 1.90 | Yes | Yes | No |
| 206780_at | GAD2 | 9.19E-02 | 0.25929 | 1.14 | 1.92E-04 | 0.04434 | 1.33 | 3.34E-02 | 0.47 | 1.17 | Yes | Yes | No |
| 212815_at | ASCC3 | 1.79E-05 | 0.01572 | 1.58 | 1.98E-04 | 0.04518 | 1.44 | 3.51E-01 | 0.71 | -1.09 | Yes | Yes | Yes |
| 211297_s_at | CDK7 | 1.05E-04 | 0.02572 | 1.51 | 2.00E-04 | 0.04540 | 1.45 | 6.97E-01 | 0.82 | -1.04 | Yes | Yes | Yes |
| 226419_s_at | FLJ44342 | 1.10E-01 | 0.27938 | 1.14 | 2.05E-04 | 0.04540 | 1.38 | 2.84E-02 | 0.46 | 1.20 | No | Yes | No |
| 202959_at | MUT | 4.06E-03 | 0.07875 | 1.27 | 2.08E-04 | 0.04555 | 1.36 | 4.03E-01 | 0.73 | 1.07 | No | Yes | No |
| 228275_at | LOC100505687 | 2.62E-04 | 0.03179 | 1.45 | 2.34E-04 | 0.04660 | 1.43 | 8.88E-01 | 0.85 | -1.01 | No | Yes | Yes |
| 219037_at | RRP15 | 1.08E-04 | 0.02587 | 1.63 | 2.65E-04 | 0.04660 | 1.55 | 6.41E-01 | 0.80 | -1.05 | Yes | Yes | Yes |
| 202232_s_at | EIF3M | 5.38E-04 | 0.03901 | 1.30 | 2.54E-04 | 0.04660 | 1.30 | 9.44E-01 | 0.86 | 1.00 | Yes | Yes | Yes |
| 244370_at | KIAA2022 | 5.86E-02 | 0.21825 | 1.17 | 2.46E-04 | 0.04660 | 1.37 | 6.43E-02 | 0.52 | 1.17 | No | Yes | No |
| 224759_s_at | C12orf23 | 5.96E-02 | 0.21949 | 1.17 | 2.70E-04 | 0.04660 | 1.37 | 6.74E-02 | 0.53 | 1.17 | No | Yes | No |
| 56256_at | SIDT2 | 1.28E-03 | 0.05364 | 1.28 | 2.28E-04 | 0.04660 | 1.31 | 6.89E-01 | 0.81 | 1.03 | Yes | Yes | No |
| 227876_at | ARHGAP39 | 1.63E-03 | 0.05756 | -1.34 | 2.48E-04 | 0.04660 | -1.39 | 6.48E-01 | 0.81 | -1.04 | Yes | Yes | No |
| 214769_at | CLCN4 | 2.13E-03 | 0.06260 | 1.32 | 2.60E-04 | 0.04660 | 1.38 | 5.92E-01 | 0.79 | 1.05 | Yes | Yes | No |
| 212335_at | GNS | 1.01E-02 | 0.10951 | 1.42 | 2.61E-04 | 0.04660 | 1.64 | 2.68E-01 | 0.67 | 1.16 | Yes | Yes | No |
| 236635_at | ZNF667 | 1.30E-02 | 0.12129 | 1.25 | 2.86E-04 | 0.04660 | 1.38 | 2.39E-01 | 0.65 | 1.11 | Yes | Yes | No |
| 227600_at | MRPS30 | 3.06E-02 | 0.16812 | 1.24 | 2.37E-04 | 0.04660 | 1.44 | 1.14E-01 | 0.58 | 1.16 | Yes | Yes | No |
| 236538_at | GRIA2 | 5.65E-02 | 0.21510 | 1.16 | 2.52E-04 | 0.04660 | 1.33 | 6.79E-02 | 0.53 | 1.15 | Yes | Yes | No |
| 223248_at | HSDL1 | 6.90E-02 | 0.23202 | 1.14 | 2.22E-04 | 0.04660 | 1.32 | 5.08E-02 | 0.50 | 1.15 | Yes | Yes | No |
| 226145_s_at | FRAS1 | 1.19E-01 | 0.28786 | 1.21 | 2.68E-04 | 0.04660 | 1.58 | 3.13E-02 | 0.47 | 1.30 | Yes | Yes | No |
| 228217_s_at | PSMG4 | 1.26E-01 | 0.29566 | 1.17 | 2.51E-04 | 0.04660 | 1.48 | 2.76E-02 | 0.46 | 1.26 | Yes | Yes | No |
| 231726_at | PCDHB14 | 1.69E-01 | 0.33254 | 1.11 | 2.18E-04 | 0.04660 | 1.34 | 1.68E-02 | 0.43 | 1.20 | Yes | Yes | No |
| 209533_s_at | PLAA | 1.82E-01 | 0.34326 | 1.13 | 2.30E-04 | 0.04660 | 1.42 | 1.58E-02 | 0.42 | 1.26 | Yes | Yes | No |
| 208841_s_at | G3BP2 | 1.87E-01 | 0.34654 | 1.11 | 2.86E-04 | 0.04660 | 1.34 | 1.80E-02 | 0.43 | 1.21 | Yes | Yes | No |
| 203414_at | MMD | 1.92E-01 | 0.35087 | 1.10 | 2.28E-04 | 0.04660 | 1.32 | 1.45E-02 | 0.42 | 1.20 | Yes | Yes | No |
| 1568612_at | GABRG2 | 9.42E-01 | 0.65815 | 1.01 | 2.69E-04 | 0.04660 | 1.40 | 4.96E-04 | 0.27 | 1.39 | Yes | Yes | No |
| 206976_s_at | HSPH1 | 1.51E-01 | 0.31797 | 1.11 | 2.94E-04 | 0.04751 | 1.32 | 2.46E-02 | 0.45 | 1.18 | Yes | Yes | No |
| 222824_at | NUDT5 | 1.16E-02 | 0.11560 | 1.29 | 3.01E-04 | 0.04793 | 1.44 | 2.63E-01 | 0.66 | 1.11 | Yes | Yes | No |
| 227049_at | ZADH2 | 1.08E-01 | 0.27719 | 1.16 | 3.08E-04 | 0.04846 | 1.40 | 3.88E-02 | 0.49 | 1.21 | Yes | Yes | No |
| 224617_at | PTBP3 | 1.52E-04 | 0.02834 | 1.31 | 3.10E-04 | 0.04848 | 1.27 | 6.85E-01 | 0.81 | -1.03 | Yes | Yes | Yes |
| 209506_s_at | NR2F1 | 7.94E-01 | 0.62016 | 1.02 | 3.13E-04 | 0.04848 | -1.33 | 2.02E-04 | 0.24 | -1.35 | Yes | Yes | No |
| 211379_x_at | B3GALNT1 | 1.89E-01 | 0.34824 | 1.20 | 3.24E-04 | 0.04960 | 1.65 | 1.95E-02 | 0.44 | 1.38 | Yes | Yes | No |

**Supplementary Figure 1:**
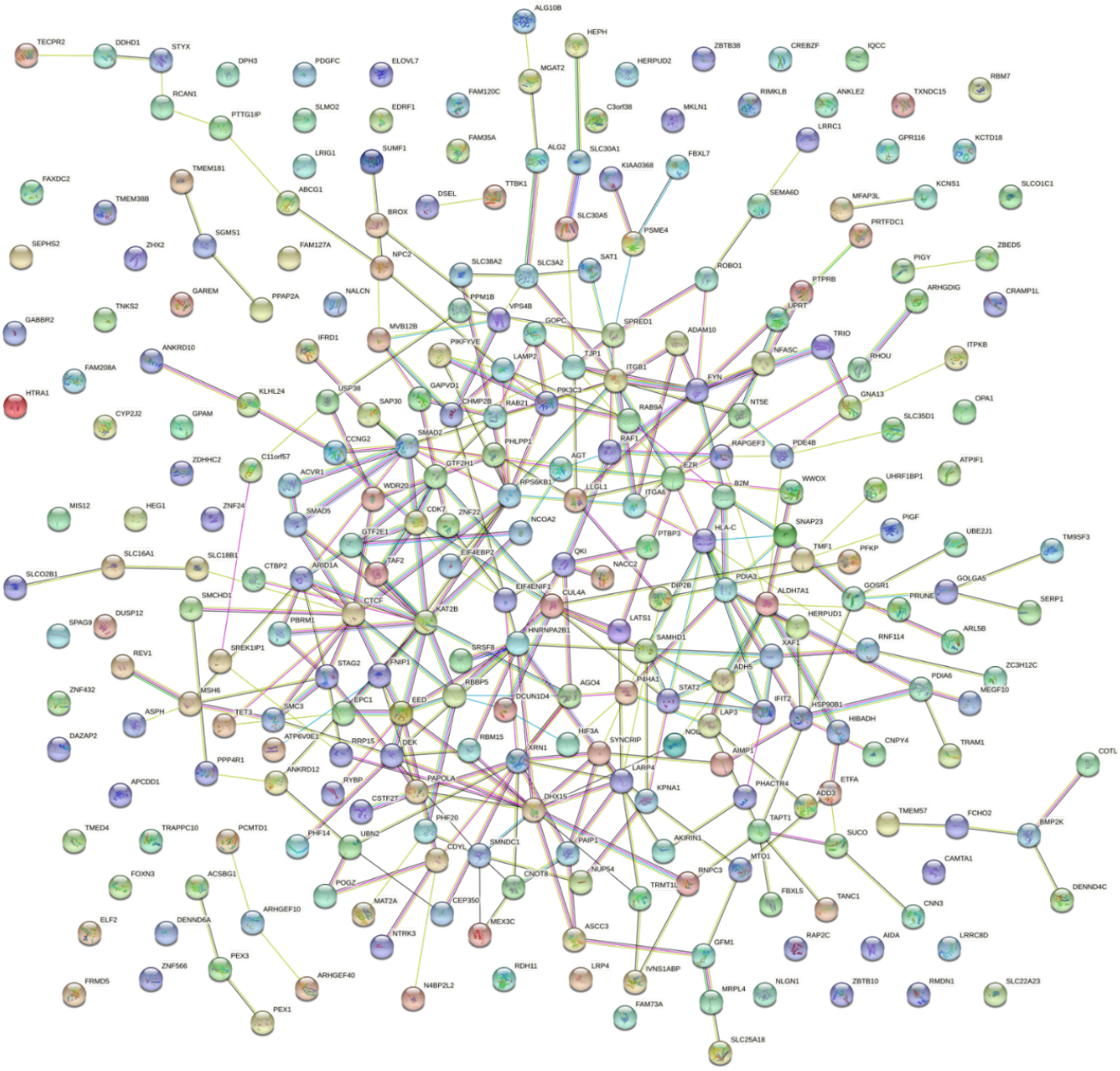
Current vs Former String network analysis.

**Supplementary Figure 2:**
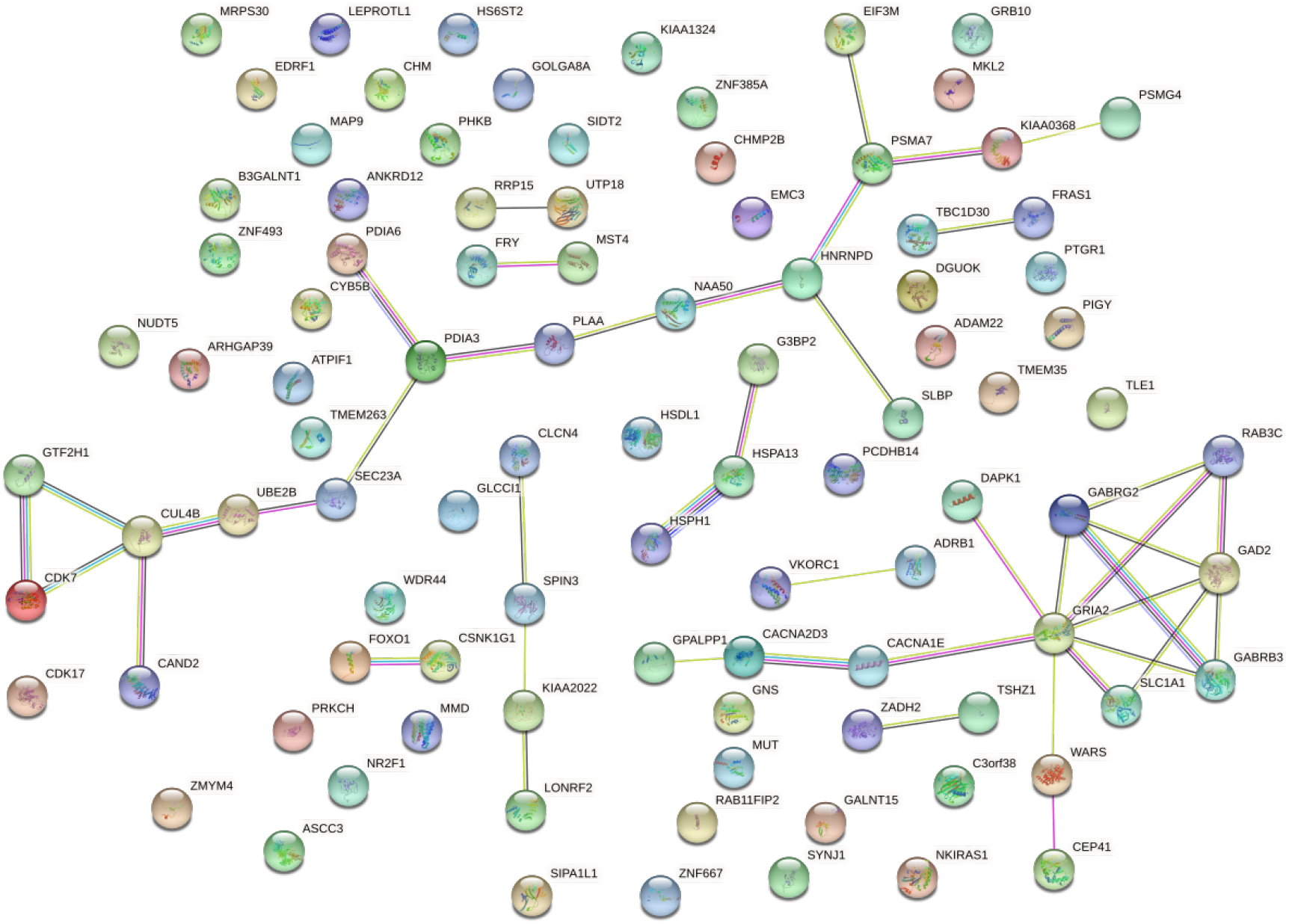
Current vs Never String network analysis.

**Supplementary Figure 3:**
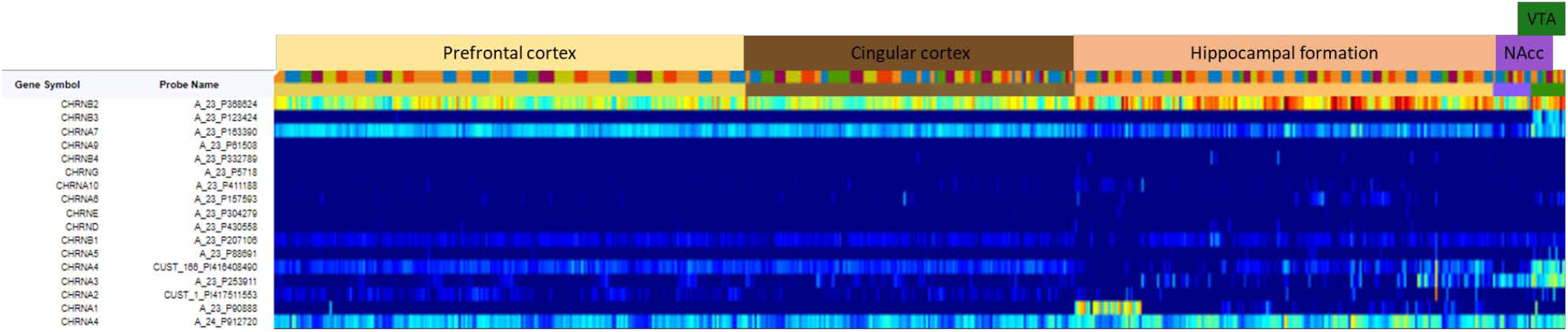
Expression of nicotinic acetylcholine receptors (nAChRs) in the human brain (data from the Allen Human Brain Atlas). Only CHRNB2, CHRNA3 and CHRNA4 receptors show detectable levels of expression in the NAcc.

**Supplementary Figure 4:**
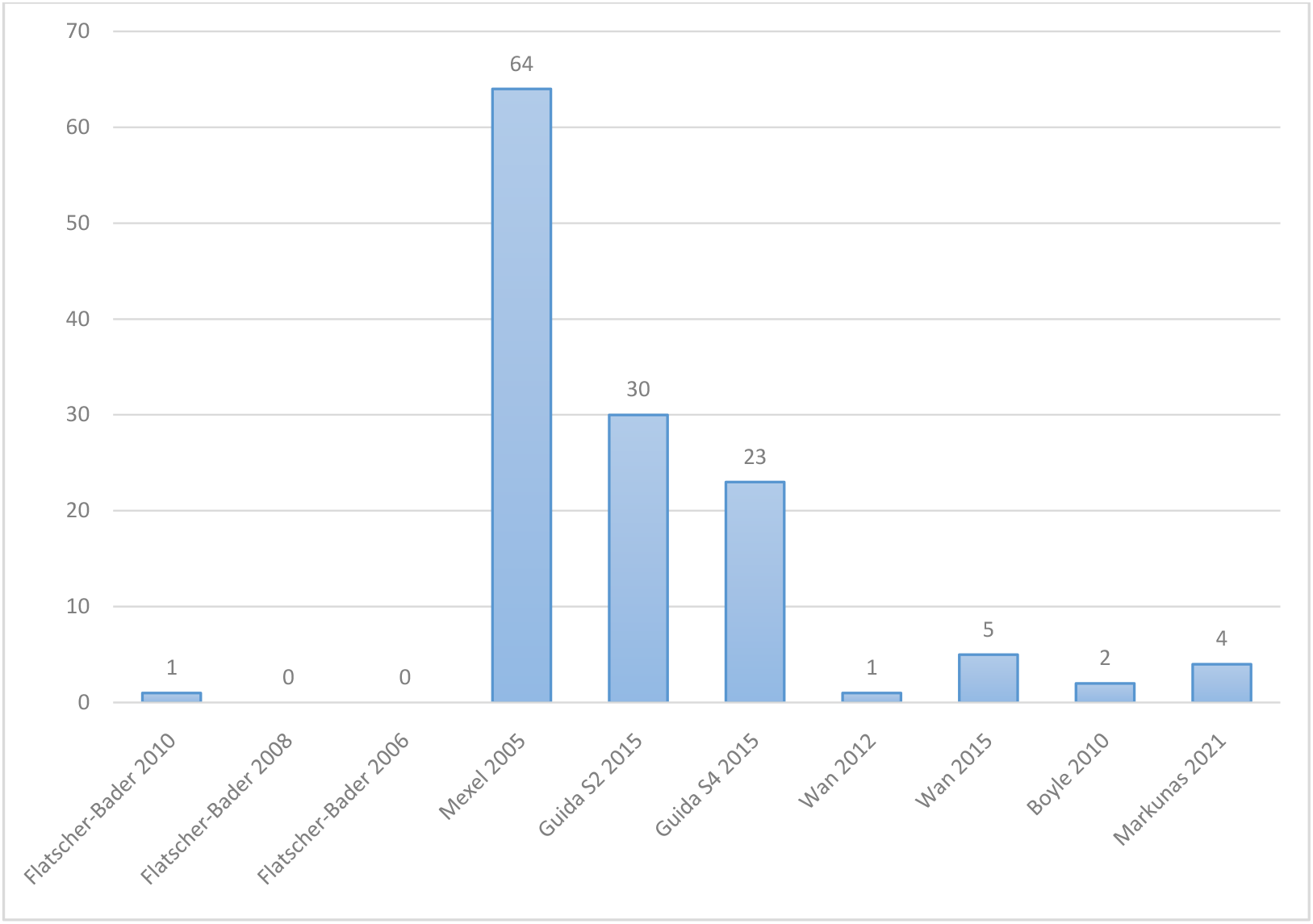
Overlap between significant differentially expressed genes found in the NAcc and previously identified smoking-associated (smokers versus non-smokers) genes looking at expression or methylation changes in the diverse tissues obtained from smokers and including buccal swabs, blood and brain tissue.

